# Brain Cell-based Genetic Subtyping and Drug Repositioning for Alzheimer Disease

**DOI:** 10.1101/2024.06.21.24309255

**Authors:** Nathan Sahelijo, Priya Rajagopalan, Lu Qian, Rufuto Rahman, Dhawal Priyadarshi, Daniel Goldstein, Sophia I. Thomopoulos, David A. Bennett, Lindsay A. Farrer, Thor D. Stein, AI4AD Consortium, Li Shen, Heng Huang, Kwangsik Nho, Saykin J. Andrew, Christos Davatzikos, Paul M. Thompson, Julia TCW, Gyungah R. Jun

**Affiliations:** Biomedical Genetics Section, Department of Medicine, Boston University Chobanian and Avedisian School of Medicine, Boston, MA, 02118, USA; Bioinformatics Program, Faculty of Computing & Data Sciences, Boston University, Boston, MA, 02215, USA; Imaging Genetics Center, Mark & Mary Stevens Neuroimaging & Informatics Institute, Keck School of Medicine, University of Southern California, Los Angeles, CA, 90032, USA; Department of Neurology, Keck School of Medicine, University of Southern California, Los Angeles, CA, 90032, USA; Department of Pharmacology, Physiology & Biophysics, Boston University Chobanian and Avedisian School of Medicine, Boston, MA, 02118, USA; Rush Alzheimer’s Disease Center, Rush University Medical Center, Chicago, IL, 60612, USA; Department of Ophthalmology, Boston University Chobanian and Avedisian School of Medicine, Boston, MA, 02118, USA; Department of Biostatistics, Boston University School of Public Health, Boston, MA, 02118, USA; Department of Epidemiology, Boston University School of Public Health, Boston, MA, 02118, USA; Department of Neurology, Boston University Chobanian and Avedisian School of Medicine, Boston, MA, 02118, USA; Department of Pathology & Laboratory Medicine, Boston University School of Medicine, Boston, Massachusetts, 02118, USA; VA Bedford Healthcare System, Bedford, MA, 01730, USA; VA Boston Healthcare Center, Boston, MA, 02130, USA; Departments of Biostatistics, Epidemiology, and Informatics, University of Pennsylvania School of Medicine, Philadelphia, PA, 19104, USA; Department of Computer Science at the University of Maryland College Park, College Park, MD, USA; Department of Radiology and Imaging Sciences and Medical and Molecular Genetics, Indiana University School of Medicine, Indianapolis, IN, 46202, USA; Indiana Alzheimer’s Disease Research Center, Indiana University School of Medicine, Indianapolis, IN, 46202, USA; Center for Computational Biology and Bioinformatics, Indiana University School of Medicine, Indianapolis, IN, 46202, USA; Department of Radiology, University of Pennsylvania School of Medicine, Philadelphia, PA, 19104, USA

**Keywords:** Alzheimer’s disease, cell-level co-expression network, cognitive performance, precision medicine, cell-based polygenic risk score, patient stratification, PageRank algorithm, drug repositioning

## Abstract

Alzheimer’s Disease (AD) is characterized by its complex and heterogeneous etiology and gradual progression, leading to high drug failure rates in late-stage clinical trials. In order to better stratify individuals at risk for AD and discern potential therapeutic targets we employed a novel procedure utilizing cell-based co-regulated gene networks and polygenic risk scores (cbPRSs). After defining genetic subtypes using extremes of cbPRS distributions, we evaluated correlations of the genetic subtypes with previously defined AD subtypes defined on the basis of domain-specific cognitive functioning and neuroimaging biomarkers. Employing a PageRank algorithm, we identified priority gene targets for the genetic subtypes. Pathway analysis of priority genes demonstrated associations with neurodegeneration and suggested candidate drugs currently utilized in diabetes, hypertension, and epilepsy for repositioning in AD. Experimental validation utilizing human induced pluripotent stem cell (hiPSC)-derived astrocytes demonstrated the modifying effects of estradiol, levetiracetam, and pioglitazone on expression of *APOE* and complement *C4* genes, suggesting potential repositioning for AD.

## Main

Late-onset Alzheimer’s disease (AD) is a progressive neurodegenerative disorder accounting for up to 80% of all dementia cases^1^. A neuropathological diagnosis of AD is based on the accumulation of extracellular amyloid-b (Ab) plaques and intracellular neuronal tau tangles in autopsied brains, while clinically diagnosed AD is primarily characterized by episodic memory loss and impairment in executive functioning. Clinically and neuropathologically diagnosed AD patients manifest different cognitive symptoms^2^ and neuroanatomical patterns^3^ with varying disease progression rates^4,5^. The presence of diverse clinical and neuropathological presentations in AD cases poses a significant challenge to the development of effective prevention and treatment strategies. The cost associated with developing drugs for AD is a substantial burden on the pharmaceutical industry and healthcare systems, where the development of a single AD drug can cost more than $5 billion. The cumulative cost of conducting clinical trials for AD, from 1995 to the present, has been estimated to be a staggering $42.5 billion^6^. AD drugs have a 99% failure rate in clinical trials; only seven drugs have been approved by the Food and Drug Administration (FDA)^7^, some of which are no longer used. This high attrition rate underscores the urgent need for innovative and more effective approaches to address the complexity and heterogeneity of AD manifestations. A recent simulation study confirmed that patient heterogeneity in AD is a primary reason for failure in late-stage AD trials by masking trial efficacy^8^. This variability presents a substantial challenge in assessing the true impact of experimental drugs in a diverse patient population. Repositioning of drugs from other disease indications to AD is a crucial complementary strategy but heterogeneity among AD patients also hampers the success of drug repositioning efforts. Addressing this heterogeneity is a key priority in the quest for more effective AD treatments and in mitigating the financial burden associated with drug development and repositioning for AD.

Twin studies have estimated the heritability for AD to be greater than 50%^9^. A large-scale genome-wide association study (GWAS) found robust evidence for over 75 loci for AD^10^. However, none of these findings yet translated into an effective disease-modifying therapy or drug repositioning for AD, arguably due to small effect sizes and incomplete understanding of underlying disease-modifying functions of the identified variants^11^. Polygenic risk scores (PRS) serve as an efficient measure of genetic liability by summing up the effect sizes of all risk alleles^12^. A recent study demonstrated that polygenic risk scores from co-expressed networks in brain tissues explain a portion of clinical heterogeneity in AD patients^13^. Phenotypic variability in AD may be due to explained by heterogeneous disease mechanisms, especially at the brain cellular level. Genomic annotations at the single tissue level can improve our understanding on the etiology of complex human diseases including AD^14^. A previous study demonstrated that individuals carrying AD-associated genetic variants exhibit distinct patterns of cellular composition in their brains^15^. This suggests the existence of unique genetically influenced disease mechanisms across different brain cell types in the human brain, depending on the presence of specific genetic variants. A cellular-level perspective may offer crucial insights into the development of more precise and effective AD treatments designed to address the genetic and cellular characteristics of individual AD patients.

In this study, we established and validated a novel framework for cellular network-based genetic subtyping and drug repositioning for AD. First, we identified co-expressed genes in each network at the cellular level and confirmed preservation and association with AD. Second, we used AD-associated genetic variants that are enriched in brain cell type-specific networks as patient selection markers and defined low and high-risk subjects for the defined cell types (genetic subtypes) in two independent datasets. We validated differential effects between low and high-risk subjects on fluid and neuroimaging biomarker measurements and clinical progression of incident AD cases. Third, we prioritized drug targets from the genes in each cellular network using a graph-based algorithm and nominated FDA-approved drugs that modulated the expression of the prioritized targets in each cell-based network. Finally, we validated the effects of the top-ranked drugs in the designated brain cell type using human induced pluripotent stem cell (hiPSC)-derived astrocytes.

## Results

The overall study design and data sources in each phase are illustrated in **Supplementary Fig. 1**. Phase 1 of this study generated cell type-specific co-expressed gene networks. Genes from the preserved networks were selected as patient selection markers for cell-based genetic subtypes. Phase 2 established genetic subtypes using cell-based polygenic risk scores (cbPRSs) and evaluated the relationship between quantitative cbPRS and previously defined AD subgroups. Subsequently, we defined genetic subtypes by dichotomizing cbPRSs using extreme values and characterized newly defined genetic subtypes using endophenotypes and clinical progression rates of AD. Phase 3 prioritized targets of the genes from the cell-based networks using a graph-based algorithm and nominated existing drugs for repositioning in AD. Based on neuropathological characteristics of the prioritized targets and drugs, we conducted experiments using hiPSC-derived astrocytes.

### Identified Cell-based Networks

We discovered 60 co-expressed gene networks specific for brain cell types using single nuclei RNA sequencing (snRNA-seq) data from the prefrontal cortex area of 48 autopsied brains in the Religious Orders Study and Rush Memory and Aging Project (ROSMAP) with 24 AD cases and 24 controls, and validated these networks by evaluating analogous data obtained from the brains of 21 AD cases and 9 controls in the Southwest Dementia Brain Bank (SWDBB) (**Supplementary Table 1**). We identified a total of 60 co-expression networks in the discovery dataset. Due to the sparse nature of snRNA-seq data and weak average correlation, we applied a stringent z-summary cutoff of 10 or greater to ensure network reproducibility. Analysis of mean expression differences of eigengenes for each network revealed four of the preserved networks including two identified in astrocytes (Ast: M2, M10) and two identified in oligodendrocytes (Oli: M45, M50), were significantly associated with AD (P<2×10^-3^) (**Supplementary Fig. 2** and **Table 1**). All four networks were highly preserved in the validation set with a preservation (z-summary) score of at least 10 (**Supplementary Fig. 3**).

**Table 1.**
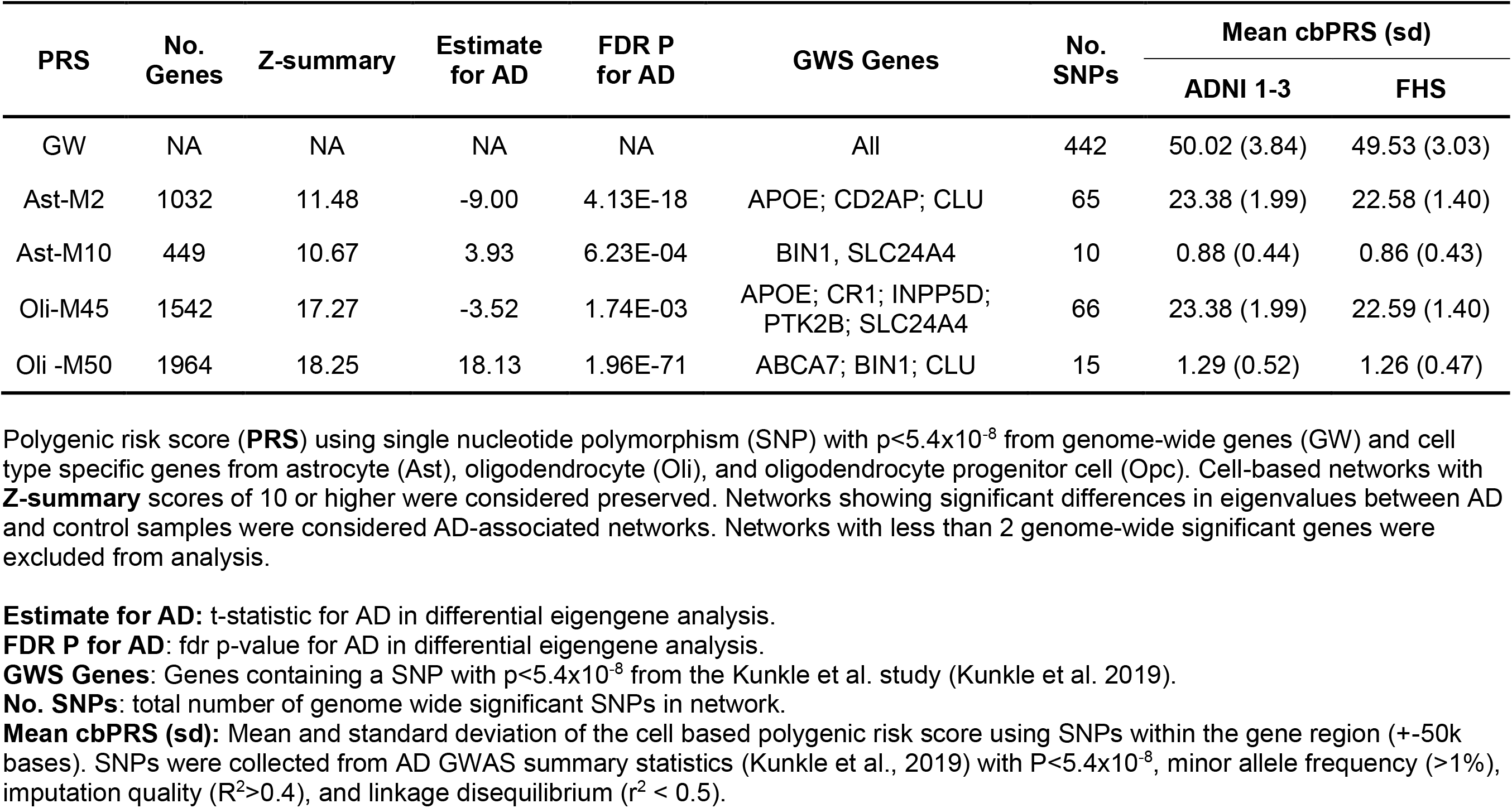
Characteristics of preserved and AD associated cell-based networks.

### Cell-based Polygenic Risk Scores for Genetic Subtypes

Polygenic risk scores were derived for 1,414 Alzheimer’s Disease Neuroimaging Initiative (ADNI; **Supplementary Table 3**) and 8,481 Framingham Heart Study (FHS; **Supplementary Table 4**) participants. The ADNI sample included 781 control (CTRL), 305 mild cognitive impairment (MCI) cases, and 328 AD cases and the FHS sample included 7,631 CTRL individuals, 310 MCI cases and 540 AD cases. The average interval between baseline and last exam was 3.45 years in ADNI and 29.46 years in FHS, indicating that the observation period was more than 9 times longer for FHS compared to ADNI participants. Cell-based polygenic risk scores (cbPRS) were calculated using the four cell-based coexpression networks as well as a genome-wide polygenic risk scores (gwPRS) for comparison. All genome-wide significant variants were included in the gwPRS in contrast to the cbPRS which included genome-wide significant variants within 50 kb of the gene loci defined by the network gene sets. All four cbPRSs as well as gwPRSs in ADNI and FHS individuals were normally distributed (**Table 1, Supplementary Table 5, and Supplementary Fig. 4**).

### Quantitative cbPRSs Explained Previously Defined AD Subgroups in ADNI

The average cbPRSs in CN, MCI, and AD cases in all ADNI subjects (ADNI1-3) for Ast-M2 and Oli-M45 were comparable to the average gwPRS (**Supplementary Fig. 5a**). We compared the association with quantitative cbPRSs on AD progression from CN to MCI, CN to AD, and MCI to AD in ADNI (**Supplementary Table 7**). The most significant association from MCI to AD in ADNI was found with gwPRS (OR=1.09; P=7.71E-05), cbPRSs from Ast-M2 (OR=1.22; P=4.41E-06), and Oli-M45 (OR=1.23; P=3.48E-06) networks (**Supplementary Fig. 5b and Supplementary Table 8**). This implied improved predictive capabilities in cbPRSs compared to gwPRS for AD progression using fewer genetic markers from cell type-specific networks.

We validated relationships between quantitative cbPRSs and previously defined cognitive subtypes in AD patients^16–19^ (**Supplementary Table 9**) and brain atrophy clusters from neuroimaging biomarker data^4^ (**Supplementary Table 10**) in ADNI 1-2/GO. Among the six cognitive subtypes for AD, quantitative cbPRSs from Ast-M2 and Oli-M45 networks were nominally significant for the AD-Memory subtype (P<0.05) with odds ratio (OR)>1 (**Supplementary Figs. 6 and 7a**). We observed significant associations after false discovery rate (FDR) adjusted P value<1.25×10^-3^ with OR>1 between quantitative cbPRSs for Ast-M2 and Oli-M45 networks and brain atrophy cluster 3 (widespread and global atrophy) and cluster 4 (localized temporal atrophy), (**Supplementary Figs. 6 and 7b**). In contrast, the quantitative cbPRSs for Ast-M2 and Oli-M45 networks were less likely members of clusters 1 or 2 (OR<1 and P<2×10^-9^).

### Cell-based Genetic Risk Status Associated Endophenotypes of AD in ADNI

We stratified low and high-risk subjects in ADNI using the first and fourth quartile of the cbPRS for each of the four networks (genetic risk status), respectively (**Supplementary Tables 11and 12**) and the FHS participants (**Supplementary Tables 13and 14**). We characterized differences of endophenotype profiles in ADNI between low- and high-risk status as a binary outcome (**Supplementary Table 15**). Risk status of Ast-M2 and Oli-M45 networks in ADNI demonstrated significant association with memory and executive function impairments, lower hippocampal volume, lower glucose metabolism, and higher global amyloid accumulation with FDR P<2.8×10^-3^ (**Fig. 1a and Supplementary Table 16**). These associations were the strongest in MCI subjects (**Table 2**). None of the endophenotypes were significantly associated with risk status of Ast-M10 and Oli-M50 networks in the total sample, CN, MCI, and AD subjects (P>0.05). The most significant association was observed for global amyloid deposition for Ast-M2 and Oli-M45 networks using MCI cases (**Table 2**). ORs for global amyloid accumulation in MCI cases were more than double those seen in AD and CN subjects (**Table 2**) suggesting a greater separation in global amyloid accumulation in the MCI subjects using the newly defined cell-based genetic risk status.

**Figure 1.**
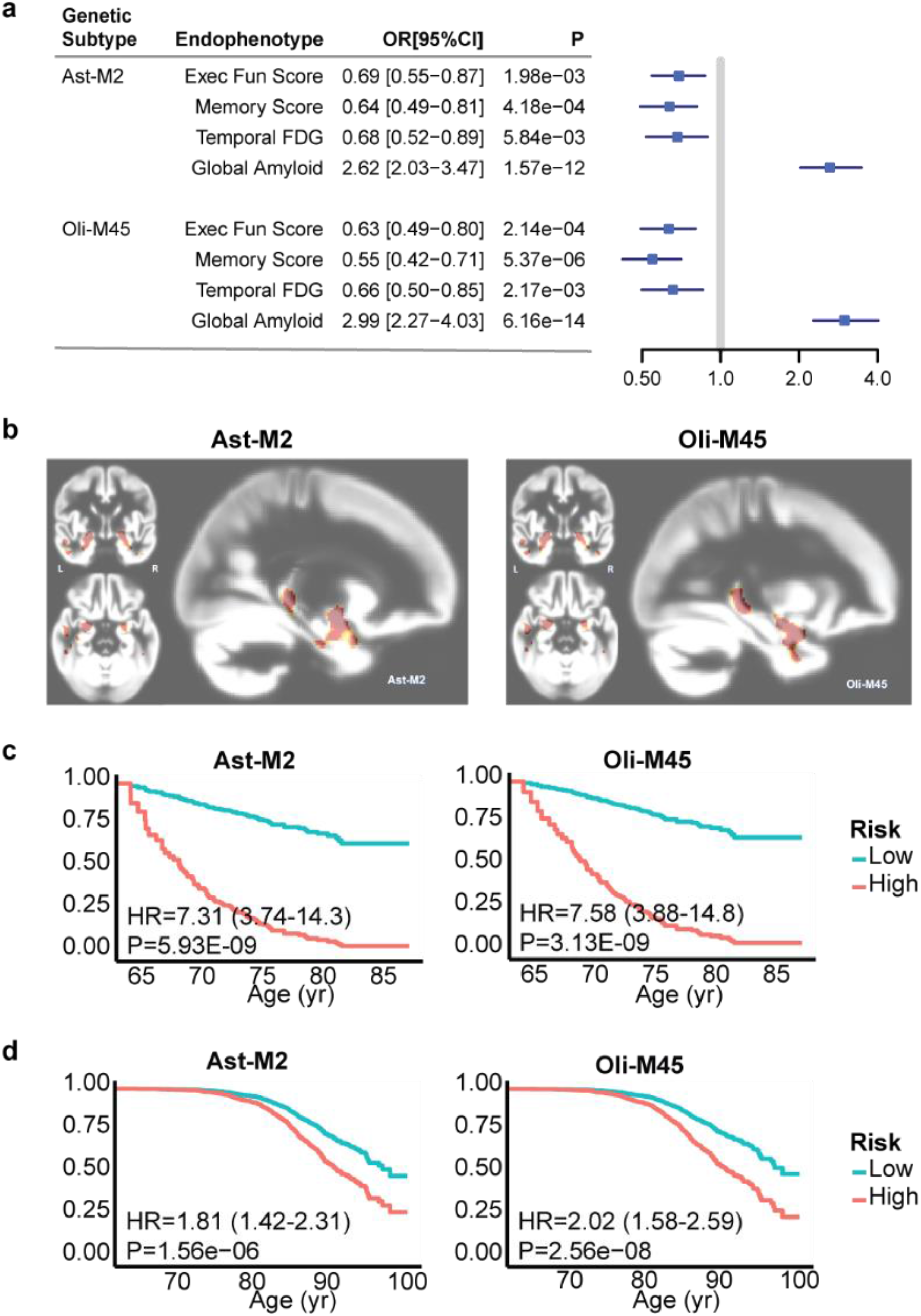
Cell-based polygenic risk scores association with AD. **a**. Association analysis of Presibo subgroups with endophenotypes of AD in ADNI 1, 2/GO, 3. Genetic subtype was treated as a binary outcome in the logistic regression model. MRI endophenotypes (hippocampal volume and entorhinal cortex thickness) include intracranial volume as a covariate. **b**. Significantly smaller brain volumes relative to the tissue probability map template predominantly in the temporal lobes, associated with Ast-M2 and Oli-M45, after correction for multiple comparisons using standard FDR at 5%. Grey matter volume deficits for Ast-M2 and Oli-M45 were detected using voxel-wise quantitative brain analysis. Regions in red highlight regions with significantly altered grey matter volumes compared to the template. Cognitive progression of Ast-M2 and Oli-M45 high and low genetic subtypes in **c**, ADNI 1, 2/GO, 3 and, **d**, FHS. All models conduct left-side censoring using a baseline age of less than 75 and a survival age greater than 65. Survival age indicates the age of earliest AD diagnosis or censoring.

**Table 2.** Association with endophenotypes of AD using genetic subtype status in CN, MCI, and AD subjects from ADNI 1-3.

| Genetic Subtype | Endophenotype | CN |  | MCI |  | AD |  |
| --- | --- | --- | --- | --- | --- | --- | --- |
|  |  | OR (95% CI) | P | OR (95% CI) | P | OR (95% CI) | P |
| Ast-M2 | Global Amyloid | 1.61 (1.07, 2.49) | 2.66E-02 | **3.72 (2.52, 5.78)** | **4.21E-10** | 1.63 (0.49, 6.84) | 4.41E-01 |
|  | Temporal FDG | 0.77 (0.4, 1.43) | 4.18E-01 | 0.69 (0.48, 0.97) | 3.65E-02 | 0.91 (0.27, 2.93) | 8.77E-01 |
|  | Memory Score | 1.28 (0.79, 2.07) | 3.15E-01 | **0.47 (0.29, 0.74)** | **1.44E-03** | 0.7 (0.12, 3.44) | 6.65E-01 |
|  | Entorhinal Cortex Thickness | 1.33 (0.81, 2.21) | 2.64E-01 | 1.11 (0.84, 1.49) | 4.54E-01 | 0.07 (0, 0.43) | 3.17E-02 |
| Oli-M45 | Global Amyloid | 1.99 (1.29, 3.19) | 2.65E-03 | **3.72 (2.5, 5.85)** | **1.12E-09** | 1.37 (0.29, 8.93) | 6.99E-01 |
|  | Temporal FDG | 0.86 (0.47, 1.52) | 6.08E-01 | 0.65 (0.45, 0.93) | 1.96E-02 | 0.92 (0.26, 3.07) | 8.94E-01 |
|  | Memory Score | 1.08 (0.67, 1.74) | 7.45E-01 | **0.41 (0.25, 0.66)** | **3.01E-04** | 3.76 (0.45, 55.18) | 2.50E-01 |
|  | Executive Function Score | 0.67 (0.41, 1.05) | 8.42E-02 | 0.68 (0.47, 0.99) | 4.48E-02 | 4.44 (1.01, 39.15) | 9.75E-02 |
|  | Entorhinal Cortex Thickness | 1.5 (0.92, 2.51) | 1.12E-01 | 1.09 (0.82, 1.47) | 5.51E-01 | 0.03 (0, 0.34) | 4.35E-02 |

**CN:** cognitively normal subjects; **MCI:** mild cognitive impairment subjects; **AD:** Alzheimer's disease cases. Odds ratio (OR) and 95% confidence interval (95% CI) were obtained from association tests of endophenotypes using genetic subtype status as a binary outcome in a logistic regression. Bold P values indicated tests passing multiple testing correction P at 2.08E-3.

Voxel based morphometry (VBM) was performed in 677 subjects (N=677; Mean age: 73.2 years) from ADNI 1, ADNI GO/2 and ADNI3. Voxel-wise gray matter volume analysis revealed that risk status for Ast-M2 and Oli-M45 were significantly associated with lower volumes predominantly in the temporal lobe, after adjusting for age, sex, intracranial volumes, and magnetic field strength. Risk status for Ast-M2 and Oli-M45 networks was significantly associated with smaller bilateral amygdala, hippocampal head and posterior body, bilateral inferior temporal gyri and left middle temporal gyrus (FDR P=0.0006 at q=0.05). Brain maps using risk status for Ast-M2 and Oli-M45 networks demonstrated significantly smaller volumes relative to the template (**Fig. 1b**).

### Cell-based Genetic Risk Status Predicted Clinical Progression to AD in ADNI and FHS

We included 477 ADNI and 540 FHS subjects that clinically progressed to AD at last exam from CNTR or MCI at baseline (**Supplementary Table 5**). We investigated differential progression rates for cell-based genetic risk status (risk status) from CNTR or MCI at baseline to exam at AD conversion in ADNI and FHS (**Supplementary Table 5**) using Cox proportional hazards models, adjusted for age at baseline, sex, and family structure (for FHS). In ADNI, risk status for Ast-M2 and Oli-M45 networks exhibited significantly different rates of progression between low- and high-risk subjects with hazard ratio (HR)>4.0, even after multiple testing correction threshold at P<3.3×10^-3^ (**Fig. 1c and Supplementary Table 17**). The pattern of findings were observed in FHS with HR>1.8 and P<10^-5^ (**Fig. 1d**), while we observed nominally significant (P<0.05) difference for risk status of Ast-M10 with HR>1.3 in FHS (**Supplementary Table 17**). The observed findings could not be fully explained by the presence of the *APOE* ε4 allele since significance for these three subtypes (Ast-M2, Ast-M10, and Oli-M45) in FHS remained nominally significant (P<0.05) in *APOE* e4 non-carriers (HR>1.4) (**Supplementary Table 18**).

### Target Prioritization for Cell-based Genetic Subtypes using the PageRank Algorithm

We analyzed the structure of the nominated networks and obtained PageRank (PR) scores of the genes within each network using the weighted PR algorithm to prioritize genes (**Fig. 2a**). PR algorithm calculated a probability distribution using the number and quality of the links between genes to quantify the importance of a node to the overall network structure. We prioritized genes for each network with positive extremes from the mean PR scores (**Fig. 2a**). This process dramatically reduced the number of genes in all networks up to 98% (**Supplementary Table 19**). We evaluated the effectiveness of PR-based prioritization using a binomial proportion test containing significantly differentially expressed genes (DEGs) between normal and AD cells in priority compared to non-priority genes. We observed increased proportion of DEGs in priority genes compared to non-priority genes with range from 16% to 62%, where the proportions of DEGs of the Ast-M2 and Oli-M50 networks were more than 50% greater in the priority compared in the non-priority genes (**Fig. 2b**). These findings suggest that PR efficiently prioritized AD-related genes while dramatically reducing the number of genes in each network.

**Figure 2.**
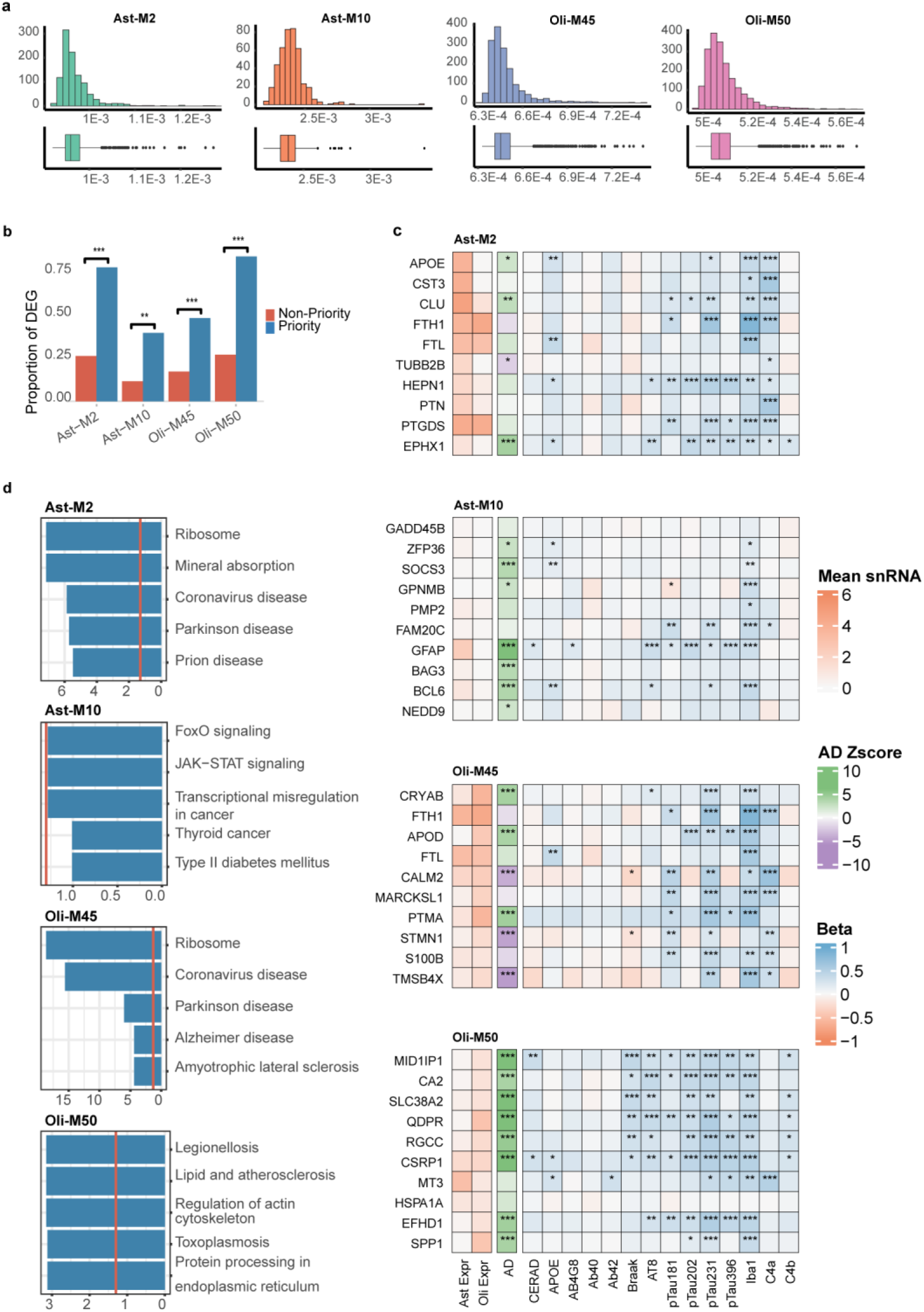
PageRank prioritization identifies differentially expressed genes (DEG). **a.** PageRank distribution for networks. Distribution displays extreme skew and highlight outlier genes with increased importance to the overall network structure. **b.** Binomial proportion tests of genes differentially expressed at the cell level in priority and non-priority gene sets. **c.** Expression profiles of the top 10 PageRank prioritized genes. Profiles display cell-specific expression and highlight distinct patterns of interactions and are ordered by decreasing rank (top to bottom). **d**. KEGG pathways enriched for priority genes.

### Molecular and Neuropathological Profiling of Priority Genes in Cell-based Networks

We confirmed expression of the top 10 ranked priority genes using the PR scores in each cell type with lower expression of the genes from the Ast-M10 network (**Fig. 2c**). Differential expression levels of the genes between AD and control brains at tissue level from the Ast-M10 and Oli-M50 were up-regulated in AD compared in control tissues (**Fig. 2c**). In contrast, the top 10 priority genes from the Ast-M2 and Oli-M45 were inconsistent in direction of tissue-based expression levels (**Fig. 2c**). Expression levels of *APOE, FTL, ZFP36, SOCS3, BCL6, CSRP1, and MT3* genes was significantly correlated with the APOE protein level in brain tissue. Most priority genes from all four networks exhibited significant associations with Iba1-positive cellular density levels (microglia marker), whereas more than 50% of the top 10 priority genes from Ast-M2 and Oli-M45 networks showed significant association with the C4a protein level (**Fig. 2c**). Priority gene profiles for the Oli-M50 network demonstrated the most significant and consistent association with Braak stage, phosphorylated Tau (pTau) levels (**Fig. 2c**). All four networks did not show a consistent relationship with amyloid pathology (**Fig. 2c**).

The top-ranked pathway using the priority genes in Ast-M2 implicated Prion disease and Parkinson’s disease (PD), while those in Oli-M45 involvled AD and amyotrophic lateral sclerosis (ALS) (**Fig. 2d**). The priority genes for both networks were enriched for PD and coronavirus disease as well as Ribosome pathway, indicating shared biological pathways. Prioritized genes for Ast-M10 networks enriched with vascular risk factors or comorbidities such as diabetes and thyroid disease however none were significant after multiple testing corrections (**Fig. 2d**). Priority genes in Oli-M50 were connected to the regulation of the actin cytoskeleton, indicating the significance in myelination and transduction mechanisms (**Fig. 2d**).

### Existing Drugs Targeting Prioritized Genes for Cell-based Genetic Subtypes

We selected drugs exhibiting the correct direction of perturbation in the expression of priority genes after treatment at the tissue level (i.e. up-regulation or down-regulation in AD compared to control tissues) (**Supplementary Tables 20-22**). The FDA-approved drugs were significantly enriched with adjusted P<0.05 using priority genes from Ast-M2 (**Supplementary Table 23**), Oli-M45 (**Supplementary Table 24**), and Oli-M50 (**Supplementary Table 25**) networks. No drugs were found with priority genes for the Ast-M10 network. We identified 54 distinct drugs for the Ast-M2, Oli-M45, and Oli-M50 networks that significantly perturbed priority genes with indications including heart disease, diabetes, inflammation, and epilepsy (**Supplementary Tables 23-25**). While most drugs (87%) perturbed all three networks, some drugs showed unique links to certain genetic subtypes. Four drugs (Atorvastatin, Dasatinib, Nicotine, and Phenol), approved for stroke, chronic myeloid leukemia, smoking cessation, and pharyngitis respectively, specifically targeted the Ast-M2 network while two drugs (Isotretinoin and Vemurafenib), approved for acne vulgaris and melanoma, altered priority genes for the Oli-M50 network (**Supplementary Tables 23-25**). Twenty-eight of the 54 candidate drugs (52%) were involved in ongoing or completed clinical trials for AD (**Supplementary Table 26**). We further examined 14 drugs targeting the Ast-M2 subtype that perturbed *APOE* expression (**Supplementary Table 26 and Supplementary Fig. 8**). Ten of the 14 drugs in clinical trials for AD (71%) are currently or previously assessed in AD clinical trials (**Supplementary Table 26**). Among these 10 drugs, we nominated four candidate drugs using the criteria of adjusted P<10^-5^ (Estradiol and Pioglitazone) as well as those used for vascular (Candesartan) or neurological (Levetiracetam) disorders (**Supplementary Table 22 and Fig. 3a**). Expression of *APOE* was increased in AD compared to control brains and positively correlated with levels of pTau231, Iba1 cell density, and complement 4A (*C4A*) in FHS (**Fig. 3b**). We investigate the effect of these four drugs and hypothesize the decreased expression of APOE and C4A levels after treatment in astrocyte cells.

**Figure 3.**
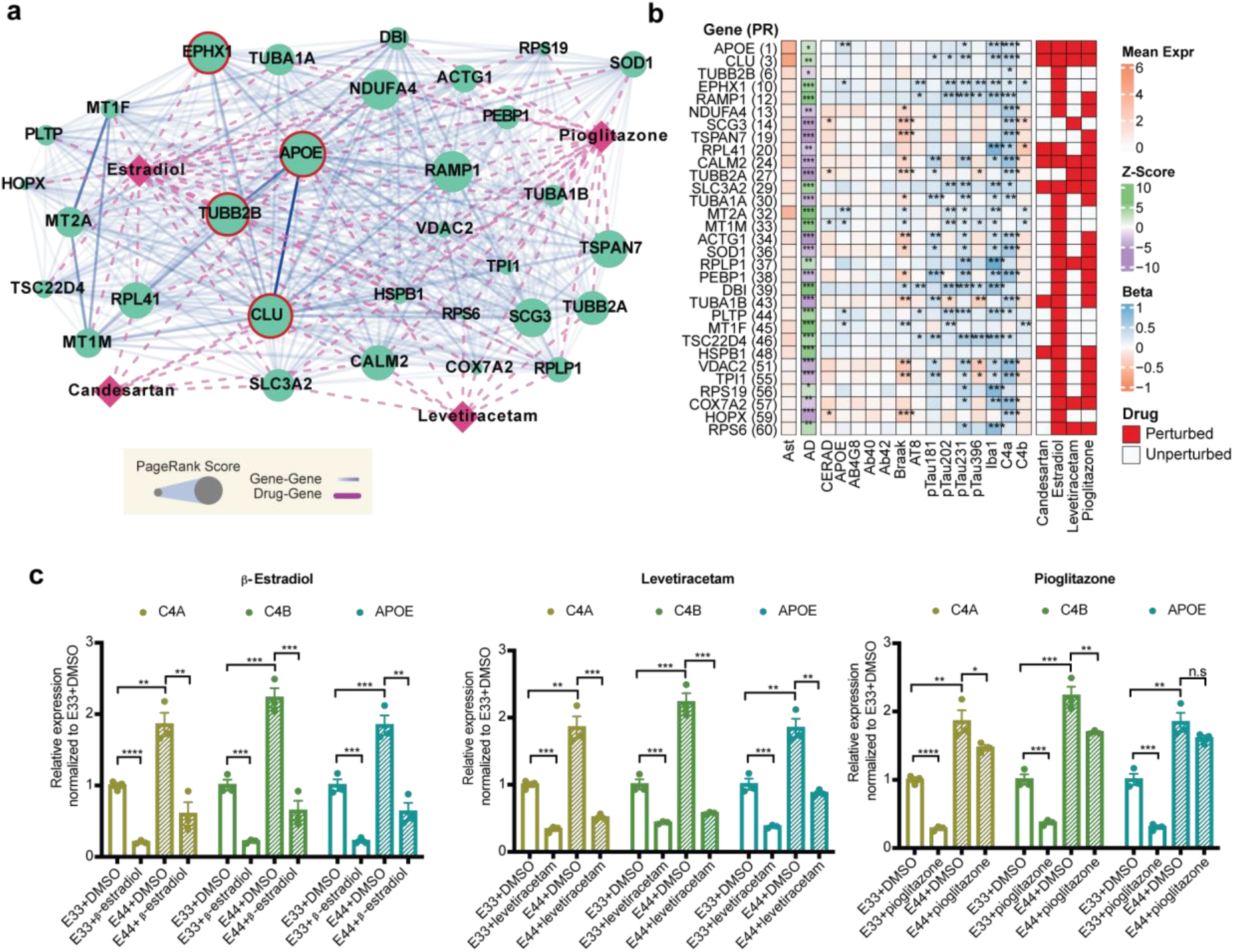
Priority genes elucidate therapeutic mechanisms perturbing network expression. **a.** Gene-drug interaction network created from priority gene targets perturbed by top-ranked drugs. Edge color indicates TOM score scaled from weakest to strongest similarity for each cell-type. Size of the node indicates network-specific PageRank rank. Genes ranked within the top 10 for Ast-M2 are outlined in red. **b.** Expression profiles of the PageRank prioritized genes perturbed by top ranked drugs. Profiles display cell-specific expression and highlight distinct biological pathways for priority genes. **c**. Measures of drug treatment efficacy in lowering *C4a* and *C4b* expression in *APOE* 33 and *APOE* 44 human iPSC-derived astrocytes. Relative expression of *C4a*, *C4b* and *APOE* genes in *APOE* 33 (E33) and *APOE* 44 (E44) iPSC-derived astrocytes treated with β-estradiol and candesartan. Gene expression was evaluated by RT-qPCR and presented as relative fold change over the DMSO vehicle in *APOE* 33 astrocytes. Error bar represented the standard error of the mean. Statistics: n=3, N=3 iPSC lines per genotype; one-tailed student t-test, *p≤0.05, **p≤0.01, ***p≤0.001, ****p≤0.0001.

### Validation of Nominated Drugs Targeting an Astrocyte Subtype using Human iPSC-Derived Astrocytes

Four FDA-approved drugs (β-Estradiol, levetiracetam, pioglitazone, and candesartan cilexetil) for the Ast-M2 subtype were administered to differentiated human induced pluripotent stem cells (hiPSCs) from a female AD patient carrying the *APOE* ε4/ε4 genotype, as well as the same cell line CRISPR-genome edited for the isogenic human *APOE* ε3/ε3 (*APOE*33) genotype. To model the findings, we used *APOE*44 AD patient carrying the AD risk SNP as well as the haplotype which mimic the risk of AD patient brain^20^. We assessed the efficacy of the drugs in lowering expression of *C4A*, *C4B*, and *APOE* in hiPSC-derived astrocytes carrying *APOE*33 and *APOE*44 genotypes after treatment of the four drugs (**Supplementary Figs. 9 and 10**). No significant cytotoxicity or compromised proliferation was observed in astrocytes after 24hrs treatment of all drugs (**Supplementary Fig. 10**). We observed significantly higher expression levels of *APOE, C4A* and *C4B* in *APOE*44 astrocytes compared in their *APOE*33 counterparts from the vehicle control (DMSO), which was consistent with the differential expression of these genes in the AD patient brains (**Fig. 3c and Supplementary Fig. 11**). Treatments of β-estradiol and levetiracetam significantly reduced the *C4A*, *C4B* and *APOE* expression (> 2-folds, p < 0.01) in both *APOE*44 and *APOE*33 hiPSC-derived astrocytes (**Fig. 3c**). After treatment of β-estradiol, *C4A*, *C4B* and *APOE* expression (> 2-folds, p < 0.01) in *APOE*33 and *APOE*44 hiPSC-derived astrocytes demonstrated 37.52% and 63.70% reduction, respectively (**Fig. 3c**). Pioglitazone also significantly decreased expression of *C4A*, *C4B* and *APOE* in *APOE*33 hiPSC-derived astrocytes, while effect of Pioglitazone was dramatically diminished in *APOE*44 hiPSC-derived astrocytes with APOE expression was not significant (**Fig. 3c**). In contrast, Candesartan exhibited the opposite direction of effect with increased expression after treatment in *APOE*33 hiPSC-derived astrocytes and did not show a significant difference in *APOE*44 hiPSC-derived astrocytes (**Supplementary Fig. 11**).

## Discussion

We developed a novel procedure using a cellular network-based approach for genetic subtyping and drug repurposing in AD. Our study implicated genetic subtypes specific to astrocyte and oligodendrocyte dysfunction. These subtypes exhibited the most significant cognitive and neuroanatomical impairments consistent with AD progression, including strong association with brain volume deficits in the hippocampus, decreased glucose metabolism, and increased global amyloid deposition. The effect of genetic subtype risk status was greatest in preclinical or MCI stages of AD progression, indicating genetically defined subtypes potentially predict which patients will transition to the clinical symptoms of AD. MCI subjects show rapid amyloid accumulation before onset of disease during the time course of AD progression^21,22^. High-risk subtypes exhibited faster rates of progression which could not be fully explained by the presence of the APOE gene. This implies cbPRSs alone can screen individuals at risk for AD and predict cellular subtypes of AD, which enables precision medicine.

Establishing robust genetic subtypes is particularly appealing for AD prevention and treatment. While genetic risk factors are commonly evaluated for association with biological pathways to understand putative links between genetic risk factors and AD pathology^10,23^, our cell-based PRSs benefit from three key improvements. First, cell specificity provides direct implications between genetic risk factors and observed cellular dysfunction. Cell-specific contributions to AD pathology are poorly understood. Recent studies looking at cell-specific perturbations to *APOE* genotypes^20,24–26^ and associations with AD biomarkers^27^ helped to elucidate our understanding of AD pathology. Most recently, cell-specific PRSs have shown to be connected to cell-type specific AD pathology^28^. Our approach is the first to provide a direct link between risk variants and cell type-specific pathology by using cell-specific coexpression networks to quantify genetic risk profiles. Second, network-specific polygenic risk scores benefit from improved power and are more discriminative than genome-wide PRS. Set-specific PRS has shown improved capabilities at explaining disease heterogeneity and is associated with distinct disease mechanisms^28–30^. Third, network structures used to identify genetic subtypes can be analyzed and interpreted to identify potential therapeutics and elucidate cellular dysfunction mechanisms. In our study, we showed the utility of the coexpression networks informing PRS to hypothesize possible disease mechanisms unique to each subtype. Experimental follow-up of prioritized network genes appears to validate network-guided PRS as a tool for hypothesis generation.

We demonstrated the potential benefit of the PageRank algorithm as a novel method to prioritize genes in networks. PageRank centrality is adept at simplifying complex interactions and identifying key nodes within dense networks. This approach successfully identified *APOE* as the top-ranked gene in the Ast-M2 network. APOE is known to disrupt the immunomodulating functions of astrocytes – the primary produces of APOE in the brain, as well as one of the major immune cells^20,31^. Expression profiles of priority genes further implicate the importance of complement-mediated neuroinflammation for the Ast-M2 and Oli-M45 subtypes. Indeed, our findings reinforce the previous reports of an *APOE* genotype-dependent mechanism linking to expression levels of *C4A* and *C4B* in AD cases^24,25^. The presence of reactive glial cells colocalizes with immune mediators like the complement system around amyloid plaques^32^. Studies suggest the complement system can interact with astrocytes through the release of pro-inflammatory cytokines leading to neuronal death and glial activation^32^. The priority genes for Ast-M10 enriched with vascular disease and neuroinflammation. Among the top-ranked priority genes for Ast-M10 was the GFAP gene (ranked 6 for the Ast-M10). GFAP is highly expressed in the brain and serves as a marker gene for astrocytes^33^ and has recently gained interest as a potential plasma biomarker capable of detecting early-stage AD^34^. GFAP plays a critical role in the activation of astrocytes and can be found highly expressed in amyloid and neuritic plaques. Increased levels of GFAP are associated with lower brain volumes^35,36^.

Oligodendrocytes play a vital role in the trophic support of axons by forming the myelin that ensheathes the CNS axon. Dysregulation of myelin can result in axonal and neuronal degeneration and lead to neuropathologies indicative of AD^37^. Among the highest ranked genes in Oli-M50, *NDRG1* (ranked 11) plays a vital role in myelinating and mature oligodendrocytes. *NDRG1* is strongly expressed in oligodendrocytes and is associated with dementia and other neuropathies^38^.

Data mining using a drug perturbation database nominated four drugs associated with the Ast-M2 subtype. Estradiol and Levetiracetam exhibited the strongest effect on APOE and C4A/C4B expression while Pioglitazone demonstrated an *APOE* genotype-dependent effect. Estradiol is an estrogen steroid hormone primarily used to treat symptoms of menopause. Studies suggest estrogen provides anti-inflammatory and neuroprotective effects^39–41^, and estrogen deficiency is associated with increased risk for AD^42^. A recent meta-analysis study found estradiol significantly reduced the risk of onset/development of AD in postmenopausal women^43^. Moreover, Estradiol upregulates AD-implicated genes including *APOE* ^44^, the top-ranked priority gene for the Ast-M2 subtype. In oligodendrocytes, estrogen induces pro-myelinating effects and promotes oligodendrocyte maturation ^45,46^. Levetiracetam is an anti-convulsant that upregulates PTGDS, prostaglandin D2 synthase (ranked 9 in Ast-M2), which regulates anti-inflammatory roles in astrocytes^47,48^ and is involved in amyloid metabolism^49^. Levetiracetam demonstrates inhibition of tau phosphorylation in rats^50^ and the amelioration of behavioral abnormalities and improved cognitive performance in mouse models^51,52^. A recent clinical trial demonstrated improved executive functioning and spatial memory among a subset of AD cases with subclinical epileptiform^53^. The results of this clinical study suggest proper subgrouping of AD cases may improve Levetiracetam’s efficacy. Pioglitazone, an anti-diabetic medication, has gained interest as an AD therapeutic by inhibiting proinflammatory genes through activation of PPARγ^54,55^. Studies confirmed anti-inflammatory effects of pioglitazone as well as improved cognitive function and decreased amyloid deposition in treated murine models of AD^54,56,57^. Early clinical results indicated improved cognitive function among diabetic patients^58^. However, additional clinical trials failed to replicate the neuroprotective effects in non-diabetic populations^59–61^.

Previous studies confirmed anti-inflammatory and neuroprotective effects through modulation of complement components for Estradiol^39,40^ and Pioglitazone^54,56,57^. The drug-induced down-regulation of *APOE* within hiPSC-derived astrocytes suggests a potential therapeutic concept for individuals at-risk for the Ast-M2 subtype, corroborating previous reports advocating estrogen therapy for *APOE-*mediated complement system perturbations^25^. Furthermore, candesartan was found to upregulate PTN (ranked 8 for Ast-M2), a key regulator of neuroinflammation^62^. As an angiotensin receptor blocker (ARB), Candesartan gained interest as a potential AD therapeutic. ARBs are thought to improve cognition by reducing inflammation^63^, decreasing oxide stress^64^, and decreasing axonal degeneration^65^. Recent clinical trials indicated improved cognitive performance in executive function tasks and episodic memory tasks in older adults with MCI^66^. A follow-up double-blind randomized placebo trial confirmed improved performance in executive function and indicated an association with decreased amyloid accumulation and increased functional network connectivity^67^.

We recognize several limitations in our study. First, the modest sample size used to generate the coexpression networks likely increased network size and made the network generation sensitive to parameter selection. Moreover, certain cell types, including Microglia, were underrepresented in the RNA dataset. Such underrepresented cell types were excluded or filtered due to low reproducibility. Additionally, the study samples were all collected from the prefrontal cortex region of the brain. AD has a predictable, region-specific impact on brain physiology and protein expression^68^. It is unclear how the region-dependent neuropathology translates to disruptions to region-specific cellular homeostasis. However, our focus on one specific brain region may have affected our ability to discern more distinct subtypes. Following studies may benefit from a variety of brain-specific samples to investigate region-specific cellular disruptions. Second, the use of AD GWAS summary statistics from studies with largely European ancestry may hinder the ability to generalize subgroups to more diverse populations. Third, the focus on genome-wide significant variants limited the variance of our PRS distributions and likely led to reduced abilities to explain AD phenotypic variance. Fourth, we conducted an undirected weighted PageRank algorithm for gene prioritization using a simple IQR cutoff to identify genes. PageRank is typically conducted on directed graphs with the undirected PageRank not exactly proportional to the degree distribution^69^. Despite the unconventional applying our IQR cutoff was able to identify genes with outsized importance in the drug network structure. Increased proportion of known AD genes within the gene subsets appears to validate our approach. Fifth, drugs identified required prior information databases in the enrichR program. While this leverages existing knowledge, it can provide a circular loop where those pathways most represented within the databases have a higher chance of being enriched. Moreover, the effect size is ignored in the drug perturbation databases casting uncertainty of the drug efficacy in perturbing network expression.

## Conclusion

We developed a novel procedure for genetic subtyping using cell-based polygenic risk scores. Applying the PageRank algorithm, we identified potential drugs for repositioning and emphasized potential biological pathways underpinning these genetic subtypes for AD. Experimental validation confirmed the perturbations of the top-ranked genes determined by the PageRank algorithm. Our study warrants the validation of our methodology in follow-up studies as well as clinical trials to facilitate precision medicine of existing drugs in the identified genetic subpopulations.

## Online Methods

### Source of Brain Cell-Level Gene Expression Data

We obtained single nuclei RNA sequencing (snRNA-seq) data from the CommonMind Consortium portal (http://www.synapse.org). The discovery set contained snRNS-seq data from the prefrontal cortex area of 48 autopsied brains with 24 AD cases and 24 controls in the Religious Orders Study and Rush Memory and Aging Project (ROSMAP) for the discovery set. All ROSMAP participants enrolled without known dementia and agreed to detailed clinical evaluation and brain donation at death. Both studies were approved by an Institutional Review Board of Rush University Medical Center. Each participant signed an informed consent, Anatomic Gift Act, and repository consent to allow their data to be repurposed. Details of the clinical and pathologic evaluation were previously reported^70–73^. For validation, we used snRNA-seq data from prefrontal cortex samples from 12 AD cases and 9 controls from the South West Dementia Brain Bank (SWDBB). Brain cell types were annotated in both the discovery and validation datasets using the expression patterns of known marker genes for astrocytes (Ast), endothelial cells (End), excitatory neurons (Ex), inhibitory neurons (In), microglia (Mic), oligodendrocytes (Oli), oligodendrocyte progenitor cells (Opc), and pericyte cells (Per)^74^. Due to the omission in the validation dataset, we excluded pericyte cells from further analysis. We reported the quality control process of the snRNA-seq data elsewhere^25^.

### Identifying Brain Cell-Level Co-Expression Networks

We generated co-expression networks with the post-QC expression data in each of seven brain cell types in the discovery set using the Weighted Gene Correlation Network Analysis (WGCNA)^75^ software tool. The WGCNA constructs co-expressed gene sets (networks) using hierarchical clustering on pairwise correlations between genes. We conducted a preservation analysis using the SWDBB dataset (validation) to ensure the reproducibility of the discovered networks. We selected preserved networks with Z-summary score from preservation analysis of 10 or more in the validation dataset^76^. Z-summary statistics compare node density and connectivity between the discovery and validation networks. The preserved networks were further evaluated for AD relevance using t-tests of mean expression differences of eigengenes from each network between AD and control cells. We considered preserved and AD-associated networks (AD-associated networks) for downstream analysis.

### Calculating Cell-Based Polygenic Risk Scores in Two Independent Studies

The Alzheimer’s Disease Neuroimaging Initiative (ADNI) is a clinical-based longitudinal study collecting clinical, imaging, genetic, and biomarker data to develop early detection mechanisms and assess the progression of AD. Genetic and phenotypic data for ADNI participants were obtained from the LONI website (http://adni.loni.usc.edu). The combined set contained 607 cognitively normal (CN), 813 mild cognitive impairment (MCI), and 516 AD subjects. Details on the imputation and quality control methods of the ADNI genetic data were previously published^13^.

The Framingham Heart Study (FHS) is a community-based longitudinal family-based study of health that surveilled participants for cognitive decline and dementia using protocols described elsewhere^77^. Since its inception in 1948, the FHS has grown to include imaging, clinical, and plasma biomarker data to study the progression of various forms of dementia. This study included 7,460 cognitively normal, 540 AD, and 481 MCI subjects where the diagnosis was defined at the last exam in 2019.

Cell-based polygenic risk scores (cbPRSs) were calculated using single nucleotide polymorphisms (SNPs) with P<5.4×10^-8^ from the previous AD GWAS^78^ that locate within 50 kb of a gene in a network. The SNPs were further excluded based on minor allele frequency (MAF)<1%, imputation quality (R^2^)<0.3 and high linkage disequilibrium (LD, r^2^>0.5). Any networks containing SNPs from only one gene or with skewed cbPRS distribution (mean cbPRS<0.05) in both studies were excluded for further analysis. We also generated genome-wide polygenic risk score (gwPRS) using all SNPs with P<5.4×10^-8^ across the genome from the previous AD GWAS.^78^ We generated cbPRSs and gwPRS for the ADNI 1-2, ADNI 3, ADNI 1-3, and FHS subjects separately.

### Correlating Cell-based and Genome-Wide Polygenic Risk Scores with Previously Defined

We conducted association of quantitative cbPRSs and gwPRS for clinical diagnosis and progression from CN to MCI, MCI to AD, and CN to AD as a binary outcome adjusting for age at last and sex based on their diagnostic status between baseline and at the last exam.

Previous studies explained clinical heterogeneity by differentiating subjects into biologically distinct cognitive subtypes in ADNI 1 and 2/GO (ADNI 1-2)^16–19^. The cognitive subtypes consisted of AD-executive function (n=16), AD-language (n=48), AD-memory (n=184), AD-multi-domain (n=25), AD-no-domain (275), and AD-visuospatial (n=85) subgroups. Another study defined image-based clusters and identified distinct brain atrophy patterns among AD and cognitively normal individuals^4^. The neuroimaging clusters assigned ADNI 1-2 subjects into one of 4 clusters^4^. Each subject was assigned to a membership of one cognitive subtype (coded as 1) without overlapping assignment across cognitive subtypes or neuroimaging clusters using the last time point. We conducted association tests with quantitative cbPRSs from an AD-associated network using a dichotomized membership status of a previously defined subgroup as a binary outcome in a logistic regression model, adjusting for age at last exam, sex, principal component (PC) 1, and PC2 as covariates. We considered P<1.25×10^-3^ for the multiple testing correction p-value (0.05/40 tests).

### Conducting Association for Genetic Risk Status with Cognitive and Neuroimaging Phenotypes in ADNI 1-3

We stratified low- and high-risk individuals for each cell type-specific network using the first and fourth quartile of each cbPRS, respectively (genetic risk status) in ADNI 1-2, ADNI 3, and the combined set of ADNI 1-3. We excluded subjects between the second and the third quartiles. We conducted association tests with cognitive and neuroimaging endophenotypes using the newly defined genetic risk status as a binary outcome in a logistic regression model. The endophenotypes included global amyloid accumulation and temporal glucose uptake from PET scans, hippocampal volume and entorhinal cortex thickness from MRI scans, and executive and memory composite scores. All endophenotypes were rank-transformed after adjusting for sex and age at exam. Models for MRI-based endophenotypes included intracranial volume as an additional covariate. The association was also evaluated for disease stage dependent effects by conducting stratified analysis separately in CN, MCI, and AD subjects defined by the last examination.

### Investigating Differential Progression Rates to AD for Genetic Risk Status in ADNI and FHS

We investigated differential progression rates from CN or MCI to AD for genetic risk status in the combined set of ADNI 1-3 (ADNI) and FHS using the Cox proportional hazard regression models in the R *survival* package. The Cox proportional hazard model for AD was compared between low and high-risk subjects for a genetic risk status. We included the interval between age at baseline and age at last exam and sex as covariates in both studies, while family structure was included as an additional covariate in the FHS. In FHS, we further evaluated effect of the *APOE* ɛ4 allele on AD progression by analyzing separately in *APOE* ɛ4 carriers and non-carriers using the same covariates.

### Imaging Methods

#### Study subjects

Magnetic resonance imaging data were obtained from the Alzheimer’s Disease Neuroimaging Initiative (ADNI) 1, 2 GO and 3. ADNI is an ongoing multisite study initially launched in 2004 by the National Institute of Health, the Food and Drug Administration, private pharmaceutical companies, and non-profit organizations for the purpose of evaluating biomarkers for AD. Written informed consent was obtained from all participants and the study was conducted according to the Good Clinical Practice guidelines, the Declaration of Helsinki, the US 21 CFR Part 50–Protection of Human Subjects, and Part 56–Institutional Review Boards. All ADNI data are publicly available and ready for downloading at https://adni.loni.usc.edu/.

In the current analysis we used MRI brain scans from a total of 677 participants from all of ADNI 1,GO, 2 and 3, across a broad cognitive spectrum including dementia (AD), mild cognitive impairment (MCI) and healthy controls (CTL). ADNI 1 (N=318; Mean age: 74.9 years) consisted of 75 AD, 160 MCI and 83 CTL participants. ADNI GO/2 (N=216; Mean age: 72.2 years) consisted of 16 AD, 141 MCI and 59 CTL participants. ADNI 3 (N=143; Mean age: 71.2 years) consisted of 6 AD, 28 MCI and 109 CTL participants.

#### Image acquisition

Our data consists of pre-existing, deidentified, three-dimensional sagittal magnetization-prepared rapid gradient-echo sequence T1-weighted brain MRI images, scanned on a 1.5 Tesla or a 3 Tesla scanner as part of the ADNI study. Details regarding the scan parameters and processing for ADNI1^79^, ADNIGO/2^80^, ADNI3^80^ are detailed previously.

#### Voxel Based Morphometry

A quantitative voxel-based analytic technique to assess the regional differences in brain volumes using voxel-based morphometry (VBM), a well-validated tool^81^ using Statistical Parametric Mapping (SPM12) software, was used to segment T1-weighted brain MRI scans as part of the ongoing ENIGMA-VBM initiative. We derived voxel-wise volumes of gray matter, white matter and cerebrospinal fluid across every single voxel in the brain using deformation mapping relative to *a priori* tissue probability map (TPM) template (Figure 2). Data analysis involved utilization of Matlab scripts and SPM12 tool (http://www.fil.ion.ucl.ac.uk./spm/) implemented in the ENIGMA-VBM pipeline^82^ (https://sites.google.com/view/enigmavbm).

### Statistical Analysis

We used a linear regression approach to assess differential voxel-wise gray matter volumes associated with the low and high-risk scores for the module based genetic risk scores after controlling for standard predictors age, sex, intracranial volume (ICV) and scanner field strength of 1.5 Tesla or 3 Tesla varying across multiple cohorts. Standard false discovery rate (FDR)^83^ was applied to correct for multiple comparisons across all gray matter voxels in the brain at the standard 5% false discovery rate (q=0.05).^13^ The critical p value, which represents the highest statistical threshold for which the statistical map controls the false discovery rate at 5%, is provided for these associations.

### Prioritizing Drug Targets within Each Cell-Based Network using the PageRank Algorithm

Considering large numbers of genes in networks, it is important to develop a robust method for prioritizing potential drug targets. We applied a weighted PageRank (PR) algorithm to calculate the gene importance to the overall network structure. We used the *igraph* R package to generate weighted PR scores for each cell-based network using the Topological Overlap Matrix similarity scores from the WGCNA as edge weights. These weights represent the likelihood of a gene interacting with another gene. We selected prioritized genes if the gene’s PR score was an outlier from the PR distribution of each cell-based network. Outliers were established using a simple interquartile range method in R. To ensure the prioritized genes were indicated with AD status we conducted a binomial proportional test comparing the proportion of AD differentially expressed genes in the priority gene set with the non-priority genes.

Priority gene sets simplified the network structure and highlighted key genes driving pathway perturbations associated with subtype etiology. We conducted pathway analysis using priority genes to identify pathways enriched with genetic subtypes. Enrichment analysis was conducted using the enrichr package^84^ in R. Priority gene sets were compared to curated pathway gene sets contained in the KEGG Human 2021 database.

### Omics Characterization of Priority Genes

We generated omics profiles for priority genes sets to characterize genetic subtypes and to supplement pathway analysis using tissue-level/cell-level DEG and neuropathological traits. The complex heatmap package^85^ in R was used to generate the combined omics heatmap profiles. The heatmaps utilized summary statistics from previous studies relating to differentially expressed genes between AD and control tissues/cells and association analysis for neuropathological traits^25^. QC and analysis methodology for these results are reported elsewhere^25^. The differential expression study used bulk RNA sequencing data from the dorsolateral prefrontal cortex area of 568 ROSMAP brains, from the temporal cortex area of 162 Mayo Clinic Study of Aging (MAYO) brains, and from the frontal cortex area of 208 the Framingham Heart Study (FHS) and Boston University Alzheimer’s Disease Center (BUADC) brains.

### Identifying Existing Drugs Targeting Cell-based Genetic Subtypes

To identify potential drugs modulating priority genes, we conducted a second enrichment analysis using the Drug_Perturbations_from_GEO_up and Drug_Perturbations_from_GEO_down databases. Since we are interested in those genes perturbed in AD, we focused our enrichment analysis on DEPs perturbed at the cell level. DEP sets were split by direction (up or down regulated) and matched with the drug database with the expected direction of effect. We obtained the list of drugs significantly modulating prioritized genes in each network with the Drug_Perturbations_from_GEO_up or Drug_Perturbations_from_GEO_down options using the *enrichrR* in the R package.

### Human iPSC-derived astrocyte culture

Pooled astrocyte lines of *APOE* 33 and *APOE* 44 genotype (*APOE* 33; n=3 & *APOE* 44; n=3) derived from hiPSCs [ref. TCW et al. Cell 2022] were cultured in Matrigel-coated 6 well plates in Astrocyte medium (ScienCell: 1801, astrocyte medium [1801-b], 2% fetal bovine serum [0010], astrocyte growth supplement [1852] and 1% Gibco™ Antibiotic-Antimycotic (100X) (15240062). Cells were seeded at densities of 3×10^^5^ cells/well of 6 well plates for the experiment and incubated in 37°C, 5% CO2 for 24 hours with the compounds in the following concentrations: Pioglitazone [50nM] (CDS021593, Sigma-Aldrich, USA), β-Estradiol [0.2 nM] (E2758, Sigma-Aldrich, USA), Levetiracetam [50 ug/ml] (PHR1447, Sigma-Aldrich, USA) and Candesartan cilexetil [5 umol/L] (SML0245, Sigma-Aldrich, USA).

### Quantitative PCR

Total RNAs from compound treated cultured cells were extracted with a RNA isolation kit (RNeasy kit; Cat. No. / ID: 74104, Qiagen, USA) following the instructions provided by the company. RNA concentration was measured spectrophotometrically using a NanoDrop 2000 (Thermo Fisher Scientific, USA). Subsequently, 1μg of total RNA was reverse transcribed to cDNA using the iScript™ Reverse Transcription Supermix (#1708840, Bio-Rad, USA). Comparative qPCR was performed in triplicates for each *APOE* 33 and *APOE* 44 cDNA sample with primers for human *C4A*, *C4B*, *APOE*, and *GAPDH* genes using Applied Biosystems™ Power SYBR™ Green PCR Master Mix (#4367659, Applied Biosystems, USA) and ran in QuantStudio™ 7 Flex Real-Time PCR System (Applied Biosystems, USA). The expression levels of target genes were normalized to the expression of respective GAPDH levels from each sample and calculated based on the comparative cycle threshold Ct method (2^-ΔΔCt^). The fold changes for each compound-treated sample were normalized to the expression levels of *APOE* 33 vehicle control (DMSO).

## Data availability

FHS data are available on the dbGaP website (https://www.ncbi.nlm.nih.gov/gap/; Study Accession ID: phs000056.v5.p3). ROSMAP resources can be requested at from the CommonMind Consortium portal (http://www.synapse.org). Data used in preparation of this article were obtained from the Alzheimer’s Disease Neuroimaging Initiative (ADNI) database (http://adni.loni.usc.edu). As such, the investigators within the ADNI contributed to the design and implementation of ADNI and/or provided data but did not participate in analysis or writing of this report. A complete listing of ADNI investigators can be found at the ADNI website (http://adni.loni.usc.edu/wp-content/uploads/how_to_apply/ADNI_Acknowledgement_List.pdf).

## Acknowledgments

This study was supported by the National Institute of Health (NIH) grants, U01AG068057, U19AG068753, U19AG079774, P30AG072978, RF1AG057519, R01AG069453, R56AG069130, U01AG032984, R01AG048927, U01AG062602, U01AG058654, RF1AG057768, and RF1AG054156. ROSMAP is supported by P30AG10161, P30AG72975, R01AG15819, R01AG17917, R01AG36042, U01AG46152, U01AG61356, R01AG082362, R56 AG078733, and K01AG062683. FHS is supported by National Heart, Lung and Blood Institute (75N92019D00031 and HHSN2682015000011). Collection of study data provided by the Rush Alzheimer’s Disease Center, Rush University Medical Center, Chicago was supported through funding by NIA grants P30-AG10161, P30-AG72975, R01-AG15819, R01-AG17917, U01-AG46152, and U01-AG61356, and funding from the Illinois Department of Public Health. We acknowledge Tony Tuck for the cell culture maintenance and support.

## Consent statement

The study protocol, design, and performance of the current study were approved by the Boston University Institutional Review Board.

## References

1. Prince, M., et al. The global impact of dementia: an analysis of prevalence, incidence, cost and trends. World Alzheimer Report 2015(2015).

2. Qiu, Y., Jacobs, D.M., Messer, K., Salmon, D.P. & Feldman, H.H. Cognitive heterogeneity in probable Alzheimer disease. Neurology 93, e778–e790 (2019).

3. Lam, B., Masellis, M., Freedman, M., Stuss, D.T. & Black, S.E. Clinical, imaging, and pathological heterogeneity of the Alzheimer’s disease syndrome. Alzheimer’s Research & Therapy 5, 1–1 (2013).

4. Dong, A., et al. Heterogeneity of neuroanatomical patterns in prodromal Alzheimer’s disease: links to cognition, progression and biomarkers. Brain, aww319–aww319 (2016).

5. Yang, Z., Wen, J. & Davatzikos, C. Smile-GANs: Semi-supervised clustering via GANs for dissecting brain disease heterogeneity from medical images. (2020).

6. Cummings, J.L., Goldman, D.P., Simmons-Stern, N.R. & Ponton, E. The costs of developing treatments for Alzheimer’s disease: A retrospective exploration. Alzheimers Dement 18, 469–477 (2022).

7. Nystuen, K.L., et al. Alzheimer’s Disease: Models and Molecular Mechanisms Informing Disease and Treatments. Bioengineering 11(2024).

8. Anderson, R.M., Hadjichrysanthou, C., Evans, S. & Wong, M.M. Why do so many clinical trials of therapies for Alzheimer’s disease fail? The Lancet 390, 2327–2329 (2017).

9. Gatz, M., et al. Role of Genes and Environments for Explaining Alzheimer Disease. Archives of General Psychiatry 63, 168–168 (2006).

10. Bellenguez, C., et al. New insights into the genetic etiology of Alzheimer’s disease and related dementias. Nat Genet 54, 412–436 (2022).

11. Tábuas-Pereira, M., Santana, I., Guerreiro, R. & Brás, J. Alzheimer’s Disease Genetics: Review of Novel Loci Associated with Disease. Current Genetic Medicine Reports 8, 1–16 (2020).

12. Lewis, C.M. & Vassos, E. Prospects for using risk scores in polygenic medicine. Genome Med 9, 96 (2017).

13. Chung, J., et al. Alzheimer’s Disease Heterogeneity Explained by Polygenic Risk Scores Derived from Brain Transcriptomic Profiles. (2022).

14. Lu, Q., et al. Systematic tissue-specific functional annotation of the human genome highlights immune-related DNA elements for late-onset Alzheimer’s disease. PLOS Genetics 13, e1006933–e1006933 (2017).

15. Li, Z., et al. Genetic variants associated with Alzheimer’s disease confer different cerebral cortex cell-type population structure. Genome Med 10, 43 (2018).

16. Mukherjee, S., et al. Genetic data and cognitively defined late-onset Alzheimer’s disease subgroups. Molecular Psychiatry 25, 2942–2951 (2020).

17. Crane, P.K., et al. Incidence of cognitively defined late-onset Alzheimer’s dementia subgroups from a prospective cohort study. Alzheimers Dement 13, 1307–1316 (2017).

18. Bauman, J., et al. Associations Between Depression, Traumatic Brain Injury, and Cognitively-Defined Late-Onset Alzheimer’s Disease Subgroups. J Alzheimers Dis 70, 611–619 (2019).

19. Groot, C., et al. Differential patterns of gray matter volumes and associated gene expression profiles in cognitively-defined Alzheimer’s disease subgroups. NeuroImage: Clinical 30, 102660–102660 (2021).

20. Tcw, J., et al. Cholesterol and matrisome pathways dysregulated in astrocytes and microglia. Cell 185, 2213–2233 e2225 (2022).

21. Moffat, G., Zhukovsky, P., Coughlan, G. & Voineskos, A.N. Unravelling the relationship between amyloid accumulation and brain network function in normal aging and very mild cognitive decline: a longitudinal analysis. Brain Commun 4, fcac282 (2022).

22. Jagust, W.J. & Landau, S.M. Temporal Dynamics of β-Amyloid Accumulation in Aging and Alzheimer Disease. Neurology 96, e1347–e1357 (2021).

23. Dourlen, P., Kilinc, D., Malmanche, N., Chapuis, J. & Lambert, J.C. The new genetic landscape of Alzheimer’s disease: from amyloid cascade to genetically driven synaptic failure hypothesis? Acta Neuropathol 138, 221–236 (2019).

24. Jun, G.R., et al. Protein phosphatase 2A and complement component 4 are linked to the protective effect of APOE varepsilon2 for Alzheimer’s disease. Alzheimers Dement 18, 2042–2054 (2022).

25. Panitch, R., et al. Integrative brain transcriptome analysis links complement component 4 and HSPA2 to the APOE ε2 protective effect in Alzheimer disease. Molecular Psychiatry 26, 6054–6064 (2021).

26. Grubman, A., et al. A single-cell atlas of entorhinal cortex from individuals with Alzheimer’s disease reveals cell-type-specific gene expression regulation. Nat Neurosci 22, 2087–2097 (2019).

27. Patel, D., et al. Cell-type-specific expression quantitative trait loci associated with Alzheimer disease in blood and brain tissue. Transl Psychiatry 11, 250 (2021).

28. Yang, H.S., et al. Cell-type-specific Alzheimer’s disease polygenic risk scores are associated with distinct disease processes in Alzheimer’s disease. Nat Commun 14, 7659 (2023).

29. Udler, M.S., et al. Type 2 diabetes genetic loci informed by multi-trait associations point to disease mechanisms and subtypes: A soft clustering analysis. PLOS Medicine 15, e1002654–e1002654 (2018).

30. Chung, J., et al. Alzheimer’s disease heterogeneity explained by polygenic risk scores derived from brain transcriptomic profiles. Alzheimers Dement 19, 5173–5184 (2023).

31. Parhizkar, S. & Holtzman, D.M. APOE mediated neuroinflammation and neurodegeneration in Alzheimer’s disease. Semin Immunol 59, 101594 (2022).

32. Shah, A., Kishore, U. & Shastri, A. Complement System in Alzheimer’s Disease. Int J Mol Sci 22(2021).

33. Yang, Z. & Wang, K.K.W. Glial fibrillary acidic protein: from intermediate filament assembly and gliosis to neurobiomarker. Trends in Neurosciences 38, 364–374 (2015).

34. Kim, K.Y., Shin, K.Y. & Chang, K.-A. GFAP as a Potential Biomarker for Alzheimer’s Disease: A Systematic Review and Meta-Analysis. Cells 12(2023).

35. Asken, B.M., et al. Lower White Matter Volume and Worse Executive Functioning Reflected in Higher Levels of Plasma GFAP among Older Adults with and Without Cognitive Impairment. Journal of the International Neuropsychological Society 28, 588–599 (2021).

36. Shir, D., et al. Association of plasma glial fibrillary acidic protein (GFAP) with neuroimaging of Alzheimer’s disease and vascular pathology. *Alzheimer’s & Dementia: Diagnosis*, Assessment & Disease Monitoring 14(2022).

37. Butt, A.M., De La Rocha, I.C. & Rivera, A. Oligodendroglial Cells in Alzheimer’s Disease. Adv Exp Med Biol 1175, 325–333 (2019).

38. Schonkeren, S.L., et al. Nervous NDRGs: the N-myc downstream-regulated gene family in the central and peripheral nervous system. Neurogenetics 20, 173–186 (2019).

39. Sarvari, M., et al. Estrogens regulate neuroinflammatory genes via estrogen receptors alpha and beta in the frontal cortex of middle-aged female rats. J Neuroinflammation 8, 82 (2011).

40. Shivers, K.Y., et al. Estrogen alters baseline and inflammatory-induced cytokine levels independent from hypothalamic-pituitary-adrenal axis activity. Cytokine 72, 121–129 (2015).

41. Giraud, S.N., Caron, C.M., Pham-Dinh, D., Kitabgi, P. & Nicot, A.B. Estradiol inhibits ongoing autoimmune neuroinflammation and NFkappaB-dependent CCL2 expression in reactive astrocytes. Proc Natl Acad Sci U S A 107, 8416–8421 (2010).

42. Barron, A.M. & Pike, C.J. Sex hormones, aging, and Alzheimer’s disease. Front Biosci (Elite Ed) 4, 976–997 (2012).

43. Song, Y.J., et al. The Effect of Estrogen Replacement Therapy on Alzheimer’s Disease and Parkinson’s Disease in Postmenopausal Women: A Meta-Analysis. Front Neurosci 14, 157 (2020).

44. Ratnakumar, A., Zimmerman, S.E., Jordan, B.A. & Mar, J.C. Estrogen activates Alzheimer’s disease genes. Alzheimers Dement (N Y) 5, 906–917 (2019).

45. Breton, J.M., Long, K.L.P., Barraza, M.K., Perloff, O.S. & Kaufer, D. Hormonal Regulation of Oligodendrogenesis II: Implications for Myelin Repair. Biomolecules 11(2021).

46. Crawford, D.K., et al. Oestrogen receptor beta ligand: a novel treatment to enhance endogenous functional remyelination. Brain 133, 2999–3016 (2010).

47. Choi, D.J., An, J., Jou, I., Park, S.M. & Joe, E.H. A Parkinson’s disease gene, DJ-1, regulates anti-inflammatory roles of astrocytes through prostaglandin D(2) synthase expression. Neurobiol Dis 127, 482–491 (2019).

48. Maesaka, J.K., et al. Prostaglandin D2 synthase: Apoptotic factor in alzheimer plasma, inducer of reactive oxygen species, inflammatory cytokines and dialysis dementia. J Nephropathol 2, 166–180 (2013).

49. Unno, K., et al. Cognitive dysfunction and amyloid β accumulation are ameliorated by the ingestion of green soybean extract in aged mice. Journal of Functional Foods 14, 345–353 (2015).

50. Alavi, M.S., Fanoudi, S., Hosseini, M. & Sadeghnia, H.R. Beneficial effects of levetiracetam in streptozotocin-induced rat model of Alzheimer’s disease. Metabolic Brain Disease 37, 689–700 (2022).

51. Sanchez, P.E., et al. Levetiracetam suppresses neuronal network dysfunction and reverses synaptic and cognitive deficits in an Alzheimer’s disease model. Proceedings of the National Academy of Sciences 109(2012).

52. Shi, J.-Q., et al. Antiepileptics Topiramate and Levetiracetam Alleviate Behavioral Deficits and Reduce Neuropathology in APPswe/PS1dE9 Transgenic Mice. CNS Neuroscience & Therapeutics 19, 871–881 (2013).

53. Vossel, K., et al. Effect of Levetiracetam on Cognition in Patients With Alzheimer Disease With and Without Epileptiform Activity. JAMA Neurology 78, 1345–1345 (2021).

54. Heneka, M.T., et al. Acute treatment with the PPARγ agonist pioglitazone and ibuprofen reduces glial inflammation and Aβ1–42 levels in APPV717I transgenic mice. Brain 128, 1442–1453 (2005).

55. Murphy, G.J. & Holder, J.C. PPAR-gamma agonists: therapeutic role in diabetes, inflammation and cancer. Trends Pharmacol Sci 21, 469–474 (2000).

56. Mandrekar-Colucci, S., Karlo, J.C. & Landreth, G.E. Mechanisms Underlying the Rapid Peroxisome Proliferator-Activated Receptor--Mediated Amyloid Clearance and Reversal of Cognitive Deficits in a Murine Model of Alzheimer’s Disease. Journal of Neuroscience 32, 10117–10128 (2012).

57. Crenshaw, D.G., et al. Effects of low doses of pioglitazone on resting-state functional connectivity in conscious rat brain. PLoS One 10, e0117973 (2015).

58. Sato, T., et al. Efficacy of PPAR-gamma agonist pioglitazone in mild Alzheimer disease. Neurobiol Aging 32, 1626–1633 (2011).

59. Geldmacher, D.S., Fritsch, T., McClendon, M.J. & Landreth, G. A randomized pilot clinical trial of the safety of pioglitazone in treatment of patients with Alzheimer disease. Arch Neurol 68, 45–50 (2011).

60. Hildreth, K.L., et al. Effects of pioglitazone or exercise in older adults with mild cognitive impairment and insulin resistance: a pilot study. Dement Geriatr Cogn Dis Extra 5, 51–63 (2015).

61. Burns, D.K., et al. The TOMMORROW study: Design of an Alzheimer’s disease delay-of-onset clinical trial. Alzheimers Dement (N Y) 5, 661–670 (2019).

62. Fernandez-Calle, R., et al. Pleiotrophin regulates microglia-mediated neuroinflammation. J Neuroinflammation 14, 46 (2017).

63. Rompe, F., et al. Direct angiotensin II type 2 receptor stimulation acts anti-inflammatory through epoxyeicosatrienoic acid and inhibition of nuclear factor kappaB. Hypertension 55, 924–931 (2010).

64. Horiuchi, M. & Mogi, M. Role of angiotensin II receptor subtype activation in cognitive function and ischaemic brain damage. Br J Pharmacol 163, 1122–1130 (2011).

65. Reinecke, K., et al. Angiotensin II accelerates functional recovery in the rat sciatic nerve in vivo: role of the AT2 receptor and the transcription factor NF-kappaB. FASEB J 17, 2094–2096 (2003).

66. Hajjar, I., et al. Effects of Candesartan vs Lisinopril on Neurocognitive Function in Older Adults With Executive Mild Cognitive Impairment: A Randomized Clinical Trial. JAMA Netw Open 3, e2012252 (2020).

67. Hajjar, I., et al. Safety and biomarker effects of candesartan in non-hypertensive adults with prodromal Alzheimer’s disease. Brain Communications 4(2022).

68. Mrdjen, D., et al. The basis of cellular and regional vulnerability in Alzheimer’s disease. Acta Neuropathol 138, 729–749 (2019).

69. Grolmusz, V. A note on the PageRank of undirected graphs. Information Processing Letters 115, 633–634 (2015).

70. Bennett, D.A., et al. Neuropathology of older persons without cognitive impairment from two community-based studies. Neurology 66, 1837–1844 (2006).

71. Bennett, D.A., et al. Natural history of mild cognitive impairment in older persons. Neurology 59, 198–205 (2002).

72. Bennett, D.A., et al. Decision rules guiding the clinical diagnosis of Alzheimer’s disease in two community-based cohort studies compared to standard practice in a clinic-based cohort study. Neuroepidemiology 27, 169–176 (2006).

73. Schneider, J.A., Arvanitakis, Z., Leurgans, S.E. & Bennett, D.A. The neuropathology of probable Alzheimer disease and mild cognitive impairment. Ann Neurol 66, 200–208 (2009).

74. Mathys, H., et al. Single-cell transcriptomic analysis of Alzheimer’s disease. Nature 570, 332–337 (2019).

## References

75. Langfelder, P. & Horvath, S. WGCNA: an R package for weighted correlation network analysis. BMC Bioinformatics 9, 559–559 (2008).

76. Langfelder, P., Luo, R., Oldham, M.C. & Horvath, S. Is My Network Module Preserved and Reproducible? PLoS Computational Biology 7, e1001057–e1001057 (2011).

77. Tsao, C.W. & Vasan, R.S. Cohort Profile: The Framingham Heart Study (FHS): overview of milestones in cardiovascular epidemiology. Int J Epidemiol 44, 1800–1813 (2015).

78. Kunkle, B.W., et al. Genetic meta-analysis of diagnosed Alzheimer’s disease identifies new risk loci and implicates Aβ, tau, immunity and lipid processing. Nature Genetics 51, 414–430 (2019).

79. Jack, C.R., Jr., et al. The Alzheimer’s Disease Neuroimaging Initiative (ADNI): MRI methods. J Magn Reson Imaging 27, 685–691 (2008).

80. Jack, C.R., Jr., et al. Magnetic resonance imaging in Alzheimer’s Disease Neuroimaging Initiative 2. Alzheimers Dement 11, 740–756 (2015).

81. Ashburner, J. & Friston, K.J. Voxel-based morphometry--the methods. Neuroimage 11, 805–821 (2000).

82. Si, S., et al. Mapping gray and white matter volume abnormalities in early-onset psychosis: an ENIGMA multicenter voxel-based morphometry study. Mol Psychiatry (2024).

83. Benjamini, Y. & Hochberg, Y. Controlling the False Discovery Rate: A Practical and Powerful Approach to Multiple Testing. Journal of the Royal Statistical Society: Series B (Methodological) 57, 289–300 (2018).

84. Chen, E.Y., et al. Enrichr: interactive and collaborative HTML5 gene list enrichment analysis tool. BMC Bioinformatics 14, 128 (2013).

85. Gu, Z., Eils, R. & Schlesner, M. Complex heatmaps reveal patterns and correlations in multidimensional genomic data. Bioinformatics 32, 2847–2849 (2016).

