## Supplemental Materials for "Brain Cell-based Genetic Subtyping and Drug Repositioning for Alzheimer Disease"

#### SUPPLENTARY MATERIALS

**SUPPLEMENTARY TABLE 1.**

**Supplementary Table 1.**
Cell count from single-nuclei RNA sequencing data.

| Cell Type | ROSMAP | | | SWDBB | | |
|---|---|---|---|---|---|---|
| | Total | AD | CN | Total | AD | CN |
| Ast | 3392 | 1830 | 1562 | 16095 | 7547 | 8548 |
| End | 116 | 56 | 60 | 1810 | 462 | 1348 |
| Ex | 34768 | 17822 | 16946 | 50428 | 24909 | 25519 |
| In | 7847 | 3840 | 4007 | 20233 | 10244 | 9989 |
| Mic | 1000 | 489 | 511 | 6285 | 3112 | 3173 |
| Oli | 18235 | 9035 | 9200 | 38985 | 17162 | 21823 |
| Opc | 2626 | 1289 | 1337 | 10128 | 4931 | 5197 |

Cell count demography from the Religious Order Study Memory and Aging Project Cell (ROSMAP) and South-West Dementia Brain Bank (SWDBB) databases for astrocytes (Ast), endothelial (End), excitatory neuron (Ex), inhibitory neuron (In), microglia (Mic), oligodendrocytes (Oli) and oligodendrocyte progenitor cells (Opc). Both datasets consist of prefrontal cortex samples with ROSMAP consisting of 24 AD and 24 control and SWDBB consisting of 12 AD and 9 control subjects. The table shows the cell count (N) in total, Alzheimer's Disease (**AD**) cases and cognitively normal controls (**CN**).

**Supplementary Table 2.**
Networks preserved at the z-summary > 5 and AD-associated cell-based networks.

| Network | Z-summary | No. Genes | FDR P for AD |
|---|---|---|---|
| Ast -M2 | 11.48 | 1032 | 2.30E-18 |
| Ast -M8 | 5.26 | 273 | 8.74E-04 |
| Ast -M9 | 9.28 | 1500 | 7.47E-19 |
| Ast -M10 | 10.67 | 449 | 2.23E-04 |
| Ex-M15 | 7.58 | 134 | 2.69E-91 |
| Ex-M17 | 9.24 | 5190 | 0.00E+00 |
| Ex-M18 | 8.75 | 46 | 1.35E-07 |
| Oli-M45 | 17.27 | 1542 | 6.96E-04 |
| Oli-M46 | 6.74 | 548 | 2.89E-14 |
| Oli-M50 | 18.25 | 1964 | 1.12E-71 |
| Oli-M51 | 8.55 | 532 | 1.55E-10 |
| Opc-M53 | 6.23 | 1087 | 7.66E-07 |

Networks have a **Z-summary** scores of 5 or higher. Networks showing significant differences in eigenvalues between AD and control samples were considered AD-associated networks. **FDR P for AD**: fdr p-value for AD in differential eigengene analysis.

**Supplementary Table 3.**
Sample size with GWAS data in ADNI.

| DX Last | ADNI 1-2/GO | | | | ADNI 3 | | | | ADNI 1-3 | | | |
|---|---|---|---|---|---|---|---|---|---|---|---|---|
| | N | N with ε4+ | % Female | Mean Age at Last (sd) | N | N with ε4+ | % Female | Mean Age at Last (sd) | N | N with ε4+ | % Female | Mean Age at Last (sd) |
| AD | 310 | 205 | 44.80 | 78.10 (7.77) | 18 | 14 | 38.90 | 74.45(9.31) | 328 | 219 | 44.5 | 77.9 (7.89) |
| CTRL | 557 | 212 | 42.00 | 80.10 (7.34) | 224 | 75 | 57.10 | 72.20 (6.05) | 781 | 287 | 46.4 | 77.8 (7.85) |
| MCI | 256 | 100 | 40.60 | 78.50 (7.89) | 49 | 22 | 34.70 | 74.30 (8.37) | 305 | 122 | 39.7 | 77.8 (8.1) |

**ADNI:** the Alzheimer's Disease Neuroimaging Initiative (ADNI); **DX Last**: AD diagnosis at last exam; **AD**: Alzheimer's disease subjects; **CTRL**: control subjects not diagnosed with AD or MCI; **MCI**: mild cognitive impairment subjects; **N**: number of subjects; **ε4+**: **ε4** carriers; % **Female**: percentage of female. **Mean Age at Last**: mean (standard deviation, sd) age at last exam.

**Supplementary Table 4.**
Sample size with GWAS data in FHS.

| Dx Last | N | N with ε4+ | % Female | Mean Age at Last (sd) |
|---|---|---|---|---|
| AD | 540 | 170 | 32.78 | 84.37 (7.56) |
| CTRL | 7631 | 1530 | 46.80 | 65.57 (14.46) |
| MCI | 310 | 76 | 45.81 | 77.38 (11.25) |

**FHS**: Framingham Heart Study; **DX Last**: AD diagnosis at last exam; **AD**: Alzheimer's disease subjects; **CTRL**: control subjects with no AD dementia or MCI; **MCI**: mild cognitive impairment subjects; **N**: number of subjects; **ε4+**: **ε4** carriers; % **Female**: percentage of female. **Mean Age at Last**: mean (standard deviation, sd) age at last exam.

**Supplementary Table 5.**
Sample with longitudinal data at baseline and at last exam in ADNI 1-3 and FHS.

| Characteristics | ADNI 1-2 | | | ADNI 3 | | | ADNI 1-3 | | | FHS | | |
|---|---|---|---|---|---|---|---|---|---|---|---|---|
| | CN | MCI | AD | CN | MCI | AD | CN | MCI | AD | CN | MCI | AD |
| N at Baseline | 315 | 610 | 197 | 210 | 62 | 18 | 525 | 672 | 215 | 8,310 | 0 | 0 |
| N at Last Exam | 284 | 369 | 456 | 209 | 59 | 21 | 493 | 428 | 477 | 7460 | 310 | 540 |
| Mean Age at Baseline (sd) | 75.29 (5.29) | 73.41 (7.52) | 75.38 (8.05) | 70.8 (5.94) | 71.83 (8.16) | 73.11 (10.63) | 73.49 (5.97) | 73.27 (7.59) | 75.19 (8.29) | 39.49 (7.1) | 39.45 (7.47) | 41.29 (7.63) |
| Mean Age at Last Exam (sd) | 79.5 (6.99) | 78.51 (7.82) | 78.58 (7.68) | 71.52 (5.88) | 73.38 (8.46) | 74.45 (9.81) | 77.16 (7.46) | 77.79 (8.1) | 77.93 (7.89) | 81.05 (10.65) | 77.38 (11.25) | 84.37 (7.56) |

**ADNI:** the Alzheimer’s Disease Neuroimaging Initiative (ADNI); **FHS**: Framingham Heart Study; **AD**: Alzheimer’s disease subjects; **CN**: cognitively normal subjects; **MCI**: mild cognitive impairment subjects; **N**: number of subjects; **ε4+**: ε4 carriers; % **Female**: percentage of female.

**Supplementary Table 6.**
Distribution of cell based PRS in 1-2/GO, 3, and the combined set.

| PRS | ADNI 1-2/GO | ADNI 3 | ADNI 1-3 |
|---|---|---|---|
| GW | 50.27 (3.8) | 49.06 (3.82) | 50.02 (3.84) |
| Ast-M2 | 23.46 (1.97) | 23.06 (2.02) | 23.38 (1.99) |
| Ast-M10 | 0.87 (0.43) | 0.91 (0.45) | 0.88 (0.44) |
| Oli-M45 | 23.47 (1.97) | 23.06 (2.02) | 23.38 (1.99) |
| Oli -M50 | 1.28 (0.51) | 1.34 (0.53) | 1.29 (0.52) |

Mean and standard deviation of the cell based polygenic risk score from astrocyte (Ast), oligodendrocyte (Oli), and oligodendrocyte progenitor cell (Opc) using SNPs within the gene region (+-50k bases). SNPs were collected from AD GWAS summary statistics [1] with $P<5.4 \times 10^{-8}$, minor allele frequency (>1%), imputation quality ($R^2>0.4$), and linkage disequilibrium ($r^2 < 0.5$).

**Supplementary Table 7.**
Characterization of AD progression in ADNI 1-3.

| Progression | N | %Female | Mean Age at Last (sd) | Mean PRS (sd) | | | | |
|---|---|---|---|---|---|---|---|---|
| | | | | GW | Ast-M2 | Ast-M10 | Oli-M45 | Oli-M50 |
| CN to MCI | 35 | 40.0 | 84.87 (5.02) | 50.1 (3.8) | 23.32 (2.07) | 0.85 (0.41) | 23.29 (2.08) | 1.27 (0.54) |
| CN to AD | 15 | 60.0 | 85.94 (4.45) | 50.61 (4.38) | 23.67 (1.9) | 0.96 (0.32) | 23.6 (1.95) | 1.31 (0.49) |
| MCI to AD | 143 | 41.3 | 78.36 (7.45) | 51.07 (3.83) | 24.02 (1.9) | 0.87 (0.41) | 24.03 (1.89) | 1.3 (0.49) |

AD progression were defined using diagnosis at baseline and at last exam.

**Supplementary Table 8.**
PRS association with AD progression in ADNI 1-3.

| PRS | CN to MCI | | CN to AD | | MCI to AD | |
|---|---|---|---|---|---|---|
| | **OR (95% CI)** | **P** | **OR (95% CI)** | **P** | **OR (95% CI)** | **P** |
| GW | 1.03 (0.93, 1.13) | 5.50E-01 | 1.09 (0.94, 1.26) | 2.28E-01 | 1.09 (1.05, 1.13) | 7.71E-05 |
| Ast-M2 | 1.02 (0.84, 1.22) | 8.40E-01 | 1.18 (0.89, 1.55) | 2.30E-01 | 1.22 (1.12, 1.22) | 4.41E-06 |
| Ast-M10 | 1.01 (0.46, 2.16) | 9.87E-01 | 1.76 (0.55, 5.44) | 3.27E-01 | 0.92 (0.61, 2.16) | 6.76E-01 |
| Oli-M45 | 1.01 (0.84, 1.21) | 9.05E-01 | 1.16 (0.87, 1.52) | 2.91E-01 | 1.23 (1.12, 1.21) | 3.48E-06 |
| Oli-M50 | 1.02 (0.51, 2.01) | 9.48E-01 | 1.16 (0.4, 3.25) | 7.80E-01 | 1.03 (0.73, 2.01) | 8.78E-01 |

Progression status was defined using the diagnosis at baseline and diagnosis at last. Logistic regression using progression status as outcome was adjusted for age at last and sex.

**Supplementary Table 9.**
Sample distribution of cognitively defined AD subtypes in ADNI 1-2/GO.

| Cognitive Subtype | N | %Female | Mean Age at Last (sd) |
|---|---|---|---|
| Executive | 16 | 38.5 | 76.9 (8.0) |
| Language | 48 | 33.3 | 78.1 (6.9) |
| Memory | 184 | 43.5 | 77.8 (7.0) |
| Multiple | 25 | 40.0 | 75.7 (6.0) |
| No domain | 275 | 42.2 | 76.8 (8.0) |
| Visuospatial | 85 | 38.8 | 75.3 (9.3) |

**N**: subgroup sample size (Mukherjee et al., 2020). **Mean Age at Last**: age at last screening expressed as mean (standard deviation).

**Supplementary Table 10.**
Sample distribution of brain atrophy clusters in ADNI 1-2/GO.

| Brain Atrophy Cluster | Characteristics of brain atrophy and cognitive decline | N | %Female | Mean Age at Last (sd) |
|---|---|---|---|---|
| 1 | Mild or no atrophy; least step on cognitive decline | 738 | 46.4 | 71.8 (7.4) |
| 2 | Widespread atrophy with greater temporal involvement; steepest cognitive decline for memory and executive function decline | 561 | 41.9 | 74.4 (8.0) |
| 3 | Widespread and global atrophy; steepest for executive function decline and intermediate steep for memory decline | 490 | 45.1 | 74.9 (7.2) |
| 4 | Localized and temporal atrophy; least steep cognitive decline | 668 | 41.6 | 76.2 (7.2) |

Distribution includes total counts from AD and control subjects from the combined ADNI-1, ADNI-GO, ADNI-2 datasets. Image patterns are treated independently as individuals may transition between image atrophy patterns (Dong et al., 2016). Four distinct clusters were identified: cluster 1 exhibits normal neuroanatomical profiles with the slowest progression; cluster 2 exhibits classic AD-associated neuroanatomical and clinical profiles with the fastest rate of progression; cluster 3 exhibits diffuse atrophy with greater executive impairment; cluster 4 exhibits medial temporal lobe atrophy with steady progression. **Mean Age at Last**: age at last screening expressed as mean (standard deviation).

**Supplementary Table 11.**
Sample characteristic of genetic subtypes in ADNI 1-3.

| Network | DX Last | Low (status=0) | | | | High (Status=1) | | | |
|---|---|---|---|---|---|---|---|---|---|
| | | **N** | **Female** | **ε4+** | **Mean Age at Last (sd)** | **N** | **Female** | **ε4+** | **Mean Age at Last (sd)** |
| M2 | AD | 78 | 32.05 | 3 | 77.76 (9.22) | 183 | 35.52 | 179 | 75.58 (7.4) |
| | CN | 77 | 49.35 | 2 | 77.29 (7.02) | 41 | 48.78 | 34 | 75.81 (8.8) |
| | MCI | 87 | 40.23 | 4 | 78.45 (8.02) | 69 | 43.48 | 67 | 74.27 (7.26) |
| M10 | AD | 126 | 35.71 | 78 | 78 (7.34) | 129 | 44.96 | 83 | 75.94 (8.15) |
| | CN | 79 | 46.84 | 23 | 78.15 (7.37) | 64 | 43.75 | 24 | 74.76 (8.01) |
| | MCI | 62 | 35.48 | 21 | 79.87 (7.87) | 81 | 43.21 | 35 | 75.6 (9.21) |
| M45 | AD | 75 | 30.67 | 4 | 77.56 (9.35) | 185 | 35.14 | 181 | 75.67 (7.26) |
| | CN | 85 | 48.24 | 2 | 76.78 (7.13) | 42 | 45.24 | 34 | 76.65 (9.05) |
| | MCI | 87 | 40.23 | 2 | 78.91 (8.07) | 70 | 45.71 | 69 | 74.08 (7.27) |
| M50 | AD | 129 | 34.88 | 90 | 78.19 (6.93) | 129 | 41.86 | 85 | 76.33 (8.28) |
| | CN | 64 | 48.44 | 17 | 78.97 (6.67) | 65 | 46.15 | 23 | 75.48 (8.32) |
| | MCI | 69 | 40.58 | 29 | 79.19 (7.93) | 79 | 37.97 | 31 | 76.1 (9.1) |

ADNI subjects stratified by presibo subgroups using cbPRS**. DX Last:** last diagnosis. **N**: number of subjects with diagnosis in strata. **High**: cbPRSs in the $4^{th}$ quartile. **Low**: cbPRSs in the $1^{st}$ quartile. **Mean Age at Last**: age at last screening expressed as mean and standard deviation (sd).

**Supplementary Table 12.**
Sample distribution of genetic subtypes in ADNI 1-3.

| | Low (status=0) | | | | High (status=1) | | | |
|---|---|---|---|---|---|---|---|---|
| **Network** | **N** | **%Female** | **Mean Conversion Age (sd)** | **Mean cbPRS (sd)** | **N** | **%Female** | **Mean Conversion Age (sd)** | **Mean cbPRS (sd)** |
| Ast-M2 | 360 | 45.3 | 77.86 (8.03) | 21.13 (0.64) | 362 | 40.3 | 75.33 (7.57) | 26.09 (1.2) |
| Ast-M10 | 362 | 42.5 | 77.88 (7.7) | 0.34 (0.13) | 363 | 45.7 | 75.77 (8.39) | 1.47 (0.22) |
| Oli-M45 | 360 | 44.4 | 77.94 (8.22) | 21.14 (0.62) | 363 | 39.9 | 75.48 (7.48) | 26.09 (1.22) |
| Oli-M50 | 362 | 42 | 77.87 (7.43) | 0.67 (0.18) | 361 | 44.3 | 76.22 (8.42) | 1.98 (0.29) |

ADNI subjects stratified into presibo subgroups using cbPRS**. N**: number of subjects in strata. **High**: cbPRSs in the $4^{th}$ quartile. **Low**: cbPRSs in the $1^{st}$ quartile. **Mean conversion age**: earliest age at AD diagnosis or censoring as mean (standard deviation). Subjects with missing sex or age were omitted.

**Supplementary Table 13.**
Sample characteristics for genetic subtypes in FHS.

| Network | DX Last | Low (Status=0) | | | | High (Status=1) | | | |
|---|---|---|---|---|---|---|---|---|---|
| | | N | Female | ε4+ | Mean Aget at Last (sd) | N | Female | ε4+ | Mean Aget at Last (sd) |
| M2 | AD | 116 | 68.1 | 8 | 85.09 (6.86) | 184 | 67.39 | 107 | 83.19 (7.18) |
| | CN | 1887 | 54.05 | 95 | 65.76 (14.52) | 1805 | 52.8 | 972 | 65.69 (14.2) |
| | MCI | 62 | 54.84 | 5 | 76.09 (12.63) | 93 | 52.69 | 52 | 75.67 (12.67) |
| M10 | AD | 113 | 63.72 | 31 | 84.89 (8.2) | 143 | 67.83 | 42 | 83.52 (8.14) |
| | CN | 1880 | 53.67 | 390 | 65.45 (14.53) | 1850 | 52.76 | 377 | 65.29 (14.05) |
| | MCI | 76 | 59.21 | 20 | 77.58 (12.47) | 87 | 51.72 | 22 | 77.46 (9.86) |
| M45 | AD | 110 | 67.27 | 9 | 84.89 (6.89) | 188 | 67.02 | 108 | 83.14 (7.21) |
| | CN | 1890 | 53.44 | 90 | 66 (14.6) | 1804 | 53.05 | 972 | 65.45 (14.17) |
| | MCI | 63 | 55.56 | 6 | 75.82 (12.77) | 91 | 54.95 | 49 | 76.4 (11.92) |
| M50 | AD | 119 | 64.71 | 41 | 85.24 (8.03) | 142 | 66.9 | 40 | 83.62 (7.95) |
| | CN | 1878 | 52.93 | 390 | 65.49 (14.47) | 1858 | 52.21 | 386 | 65.55 (14.49) |
| | MCI | 78 | 62.82 | 16 | 76.8 (12.54) | 78 | 53.85 | 19 | 76.69 (10.24) |

FHS subjects stratified by presibo subgroups using cbPRS**. DX Last:** last diagnosis. **N**: number of subjects with diagnosis in strata. **High**: cbPRSs in the 4$^{th}$ quartile. **Low**: cbPRSs in the 1$^{st}$ quartile. **Mean Age at Last**: age at last screening expressed as mean and standard deviation (sd).

**Supplementary Table 14.**
Cell-based PRS distribution in genetic subtypes in FHS.

| Network | Low (status=0) | | | | High (status=1) | | | |
|---|---|---|---|---|---|---|---|---|
| | N | %Female | Mean Conversion Age (sd) | Mean cbPRS (sd) | N | %Female | Mean Conversion Age (sd) | Mean cbPRS (sd) |
| Ast-M2 | 2120 | 54.9 | 67.65 (14.92) | 21.24 (0.56) | 2120 | 53.7 | 67.76 (14.61) | 24.75 (0.99) |
| Ast-M10 | 2120 | 54.2 | 67.32 (15.05) | 0.34 (0.12) | 2120 | 53.6 | 67.31 (14.52) | 1.44 (0.23) |
| Oli-M45 | 2120 | 54.7 | 67.75 (14.91) | 21.24 (0.56) | 2120 | 54 | 67.8 (14.6) | 24.76 (0.99) |
| Oli-M50 | 2120 | 53.6 | 67.34 (14.98) | 0.68 (0.17) | 2120 | 53.3 | 67.47 (14.84) | 1.88 (0.25) |

FHS subjects stratified into presibo subgroups using cbPRS. **N**: number of subjects in strata. **High**: cbPRSs in the 4$^{th}$ quartile. **Low**: cbPRSs in the 1$^{st}$ quartile. **Mean conversion age**: earliest age at AD diagnosis or censoring as mean (standard deviation). Subjects with missing sex or age were omitted.

**Supplementary Table 15.**
Characteristics of Image and cognitive endophenotypes in ADNI-3.

| Endophenotype | N | %Female | Mean Age at Exam (sd) |
|---|---|---|---|
| Executive Function | 1604 | 44.26 | 73.45 (7.14) |
| Memory | 1604 | 44.26 | 73.45 (7.14) |
| Global Amyloid | 1017 | 45.43 | 73.68 (7.64) |
| Temporal FDG | 1200 | 43.33 | 73.25 (7.12) |
| Hippocampal Volume | 1595 | 44.20 | 73.41 (7.13) |
| Entorhinal Cortex Thickness | 1595 | 44.20 | 73.41 (7.13) |

**Supplementary Table 16.**
Association results with endophenotypes cell-based genetic risk status using all subjects in ADNI 1-3.

| Network | Endophenotype | OR (95% CI) | P |
|---|---|---|---|
| Ast-M2 | Global Amyloid | 2.62 (2.03, 3.47) | 1.57E-12 |
| | Temporal FDG | 0.68 (0.52, 0.89) | 5.84E-03 |
| | Memory Score | 0.64 (0.49, 0.81) | 4.18E-04 |
| | Executive Function Score | 0.69 (0.55, 0.87) | 1.98E-03 |
| | Hippocampal Volume | 0.86 (0.69, 1.06) | 1.64E-01 |
| | Entorhinal Cortex Thickness | 0.9 (0.73, 1.13) | 3.69E-01 |
| Ast-M10 | Global Amyloid | 1.07 (0.87, 1.32) | 5.32E-01 |
| | Temporal FDG | 0.82 (0.62, 1.08) | 1.59E-01 |
| | Memory Score | 0.87 (0.69, 1.1) | 2.35E-01 |
| | Executive Function Score | 0.88 (0.7, 1.1) | 2.58E-01 |
| | Hippocampal Volume | 0.9 (0.72, 1.12) | 3.51E-01 |
| | Entorhinal Cortex Thickness | 1.03 (0.83, 1.28) | 7.72E-01 |
| Oli-M45 | Global Amyloid | 2.99 (2.27, 4.03) | 6.16E-14 |
| | Temporal FDG | 0.66 (0.5, 0.85) | 2.17E-03 |
| | Memory Score | 0.55 (0.42, 0.71) | 5.37E-06 |
| | Executive Function Score | 0.63 (0.49, 0.8) | 2.14E-04 |
| | Hippocampal Volume | 0.85 (0.68, 1.05) | 1.35E-01 |
| | Entorhinal Cortex Thickness | 0.89 (0.71, 1.11) | 3.16E-01 |
| Oli-M50 | Global Amyloid | 1.11 (0.9, 1.37) | 3.38E-01 |
| | Temporal FDG | 0.9 (0.69, 1.17) | 4.36E-01 |
| | Memory Score | 1 (0.8, 1.25) | 9.92E-01 |
| | Executive Function Score | 1.01 (0.81, 1.27) | 9.26E-01 |
| | Hippocampal Volume | 1.02 (0.82, 1.26) | 8.82E-01 |
| | Entorhinal Cortex Thickness | 1.14 (0.92, 1.43) | 2.36E-01 |

**OR (95% CI):** odds ratio with 95% lower and upper bound. Presibo subgroup membership was treated as a binary outcome in the logistic regression model. MRI endophenotypes (hippocampal volume and entorhinal cortex thickness) included intracranial volume and magnetic field strength as covariates. Associations passing multiple testing correction are bolded $p<1.04E-3$.

**Supplementary Table 17.**
Cox proportional hazard ratio for cell-based genetic risk status in ADNI 1-3 and FHS.

| Network | ADNI | | FHS | |
|---|---|---|---|---|
| | HR (95% CI) | P | HR (95% CI) | P |
| Ast-M2 | 7.31 (3.74-14.3) | 5.93E-09 | 1.81 (1.42-2.31) | 1.56E-06 |
| Ast-M10 | 0.93 (0.57-1.53) | 7.87E-01 | 1.41 (1.10-1.80) | 7.00E-03 |
| Oli-M45 | 7.58 (3.88-14.8) | 3.13E-09 | 2.02 (1.58-2.59) | 2.56E-08 |
| Oli-M50 | 0.95 (0.6-1.51) | 8.37E-01 | 1.25 (0.98-1.59) | 6.84E-02 |

Cox Proportional Hazard Ratios with 95% confidence interval were computed between low and high-risk subjects.
**HR**: Cox Proportional Hazard Ratio.

**Supplementary Table 18.**
Cox proportional hazard ratio for genetic subtype status in FHS stratified by APOE ε4 carrier status.

| Network | ***APOE* ε4 non-carriers** | | ***APOE* ε4 carriers** | |
|---|---|---|---|---|
| | **HR (95% CI)** | **P** | **HR (95% CI)** | **P** |
| Ast-M2 | 1.42 (1.07-1.9) | 1.70E-02 | 1.18 (0.57-2.45) | 6.54E-01 |
| Ast-M10 | 1.38 (1.02-1.87) | 3.57E-02 | 1.56 (0.99-2.46) | 5.52E-02 |
| Oli-M45 | 1.68 (1.25-2.26) | 5.89E-04 | 0.94 (0.46-1.93) | 8.65E-01 |
| Oli-M50 | 1.34 (0.99-1.81) | 5.44E-02 | 1.16 (0.74-1.83) | 5.20E-01 |

Cox Proportional Hazard Ratios with 95% confidence interval computed for low and high presibo subgroups using low as the reference group and adjusting for sex. **HR**: Cox Proportional Hazard Ratio.

**Supplementary Table 19.**
Binomial proportion test of differentially expressed genes (DEG) in AD brain cells.

| Network | No. of all genes | No. of prioritized genes | % reduction after prioritization | Proportion Test | | |
|---|---|---|---|---|---|---|
| | | | | % of DEGs in priority gens | % of DEGs in non-priority genes | P |
| Ast-M2 | 1032 | 60 | 94% | 78% | 27% | 5.0E-17 |
| Ast-M10 | 449 | 10 | 98% | 40% | 12% | 2.9E-02 |
| Oli-M45 | 1542 | 115 | 93% | 49% | 17% | 1.3E-15 |
| Oli-M50 | 1963 | 98 | 95% | 85% | 27% | 6.1E-33 |

**No. DEG:** number of genes nominally differentially expressed in AD brain cells. **Prop DEG:** proportion of genes differentially expressed.

**Supplementary Table 20.**
Differentially expressed priority genes from the Ast-M2 network with P<0.05 between AD and control brain tissue (Tisuse Zscore) (excel file).

**Supplementary Table 21.**
Differentially expressed priority genes from the Oli-M45 network with P<0.05 between AD and control brain tissue (Tisuse Zscore) (excel file).

**Supplementary Table 22.**
Differentially expressed priority genes from the Oli-M50 network with P<0.05 between AD and control brain tissue (Tisuse Zscore) (excel file).

**Supplementary Table 23.**
Approved drugs with significant enrichment for Ast-M2 priority genes differentially expressed in AD brain tissue (excel).

**Supplementary Table 24.**
Approved drugs with significant enrichment for Oli-M45 priority genes differentially expressed in AD brain tissue (excel).

**Supplementary Table 25.**
Approved drugs with significant enrichment for Oli-M50 priority genes differentially expressed in AD brain tissue (excel).

**Supplementary Table 26.**
Drugs in clinical trials for AD (excel).

**Supplementary Table 27.**
Sequences of primers.

| Primer | Direction | Sequence |
|---|---|---|
| h-C4A | Forward | 5’ GCTCACAGCCTTTGTGTTG 3’ |
| | Reverse | 5’ CTGCATGCTCCTGTCTAAC 3’ |
| h-C4B | Forward | 5’ GCTCACAGCCTTTGTGTTG 3’ |
| | Reverse | 5’ CTGCATGCTCCTATGTATCAC 3’ |
| h-APOE | Forward | 5’ GAGCAGGCCCAGCAGATA 3’ |
| | Reverse | 5’ CTGCATGTCTTCCACCAGGG 3’ |
| h-GAPDH | Forward | 5’ AGGGCTGCTTTTAACTCTGGT 3’ |
| | Reverse | 5’ CCCCACTTGATTTTGGAGGGA 3’ |

**Supplementary Figure 1.**
Overall study design in three phases. Phase 1 represented the process to identify brain cell type specific co-expression networks. Phase 2 defined and characterized genetic subtypes. Phase 3 prioritized cell-based targets for drug repositioning and validated drug effects. AD: Alzheimer’s disease; snRNA: single nuclei RNA sequencing data; PRS: polygenic risk score; ADNI: Alzheimer’s Disease Neuroimaging Initiative; FHS: Framingham Heart Study.

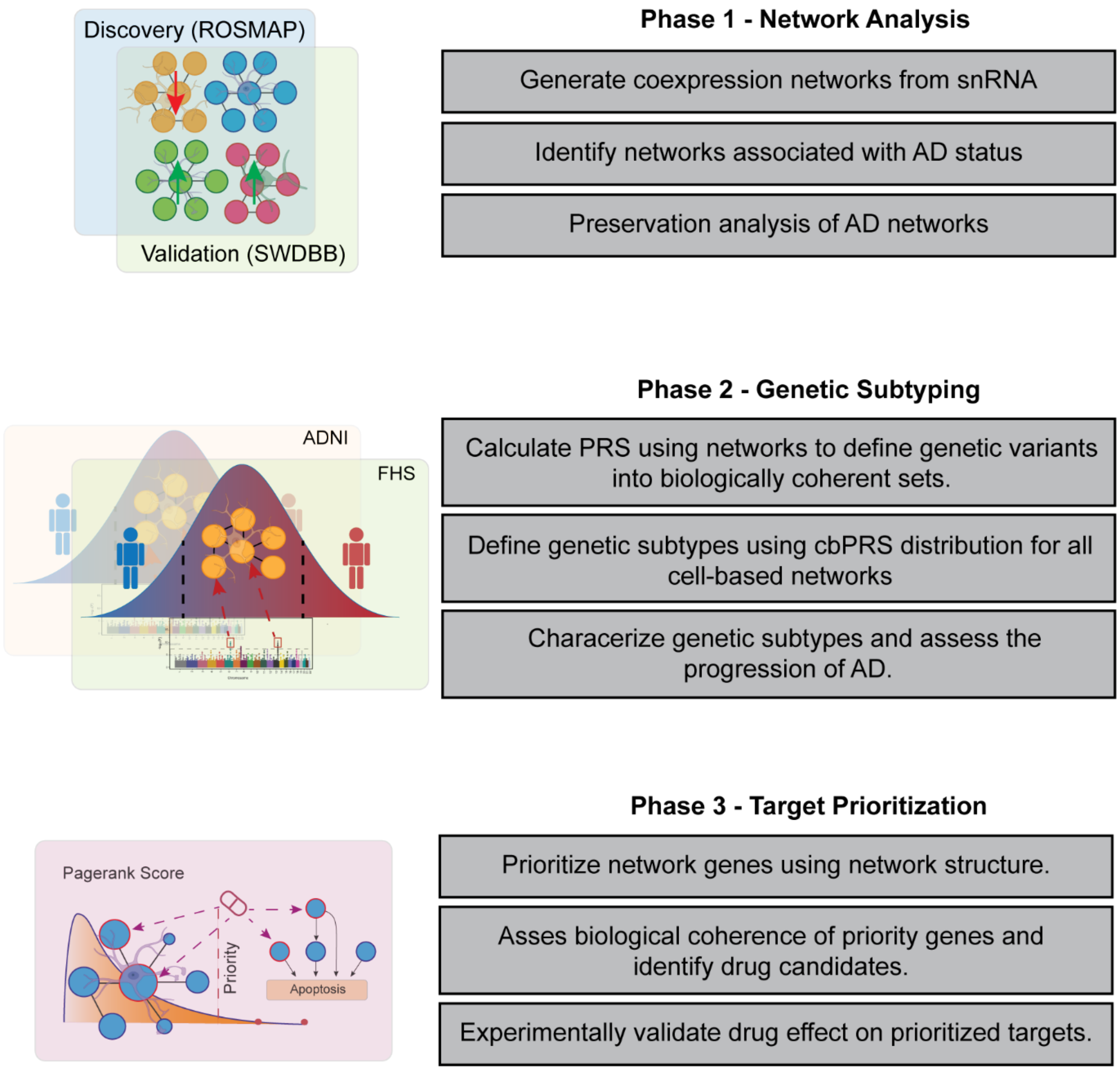

**Supplementary Figure 2.**

Eigengene expression in AD and cognitively normal cells in ROSMAP. Eigengene expression in CN and AD cells for all preserved networks.

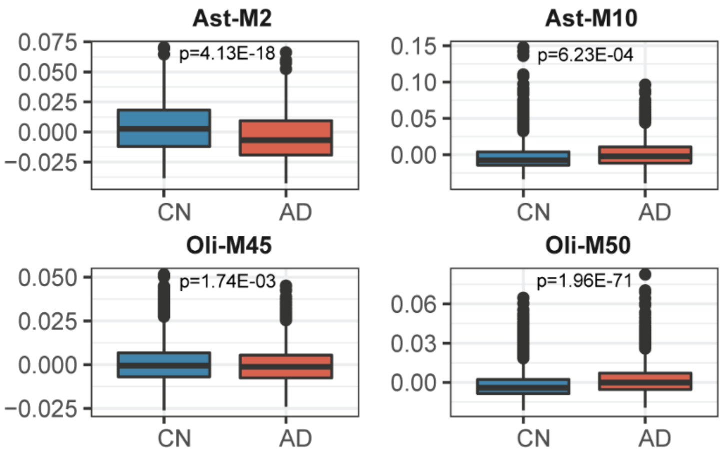

**Supplementary Figure 3.**
Preservation of WGCNA networks for Ast and Oli. Modules with a Zsummary statistic > 10 were considered preserved.

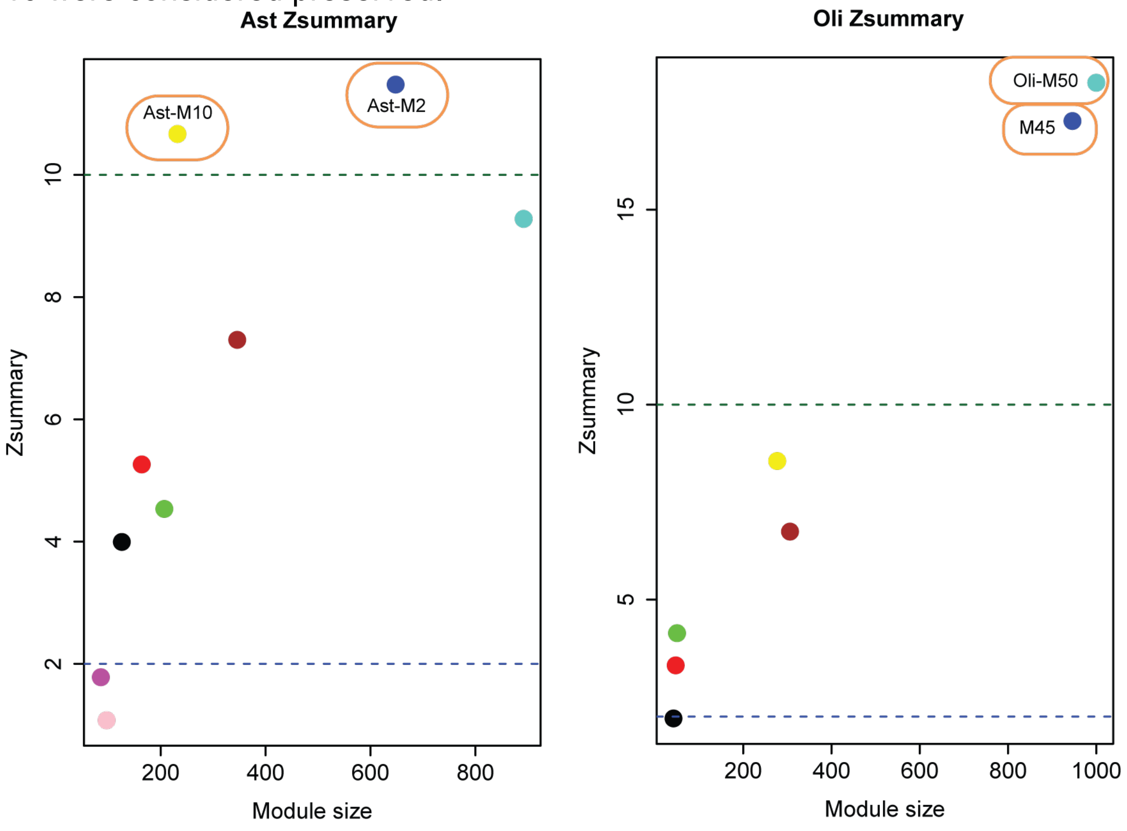

**Supplementary Figure 4.**
Distribution of quantitative cbPRSs. Relative frequency density of cbPRS scores stratified by cohort.

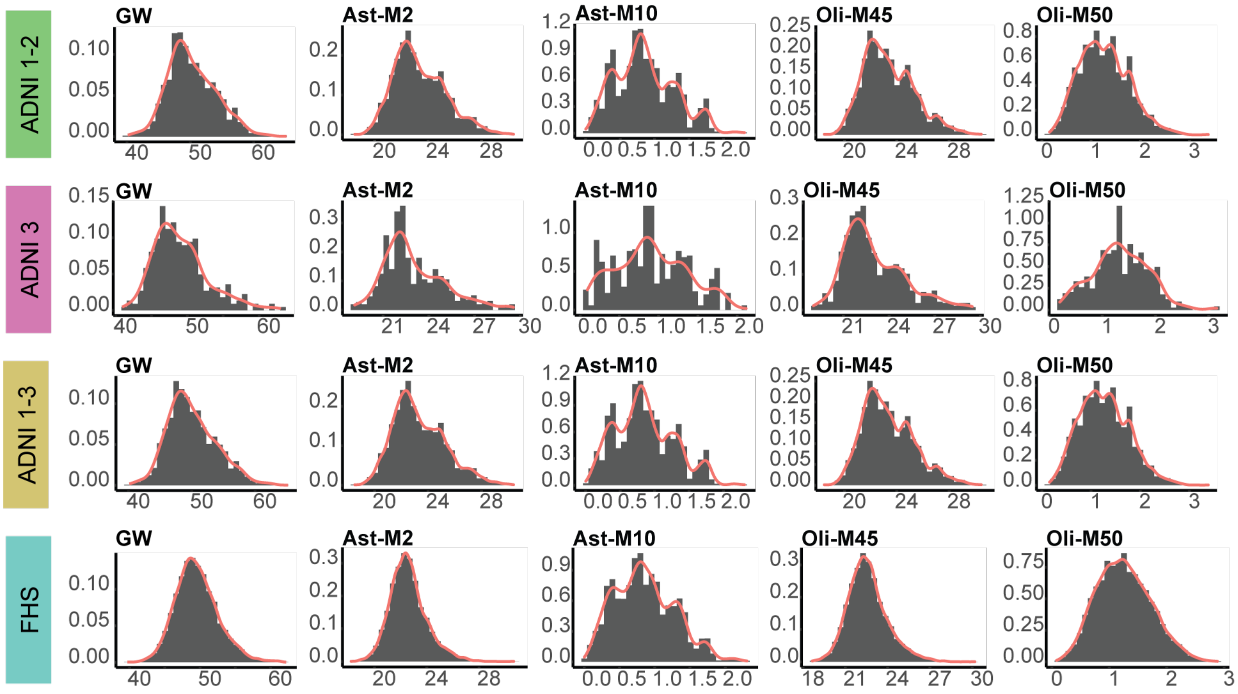

**Supplementary Figure 5.**
PRS association with cognitive diagnosis and progression in ADNI 1-3. **a.** Heatmap of mean rank-normalized genome-wide PRS (AD) and cbPRS with last diagnosis. **b.** Association of AD PRS and cbPRS with disease progression. Odds ratios (OR) were calculated using a logistic regression model adjusted for age at last and sex with disease progression as the binary outcome and raw PRS scores as predictors.

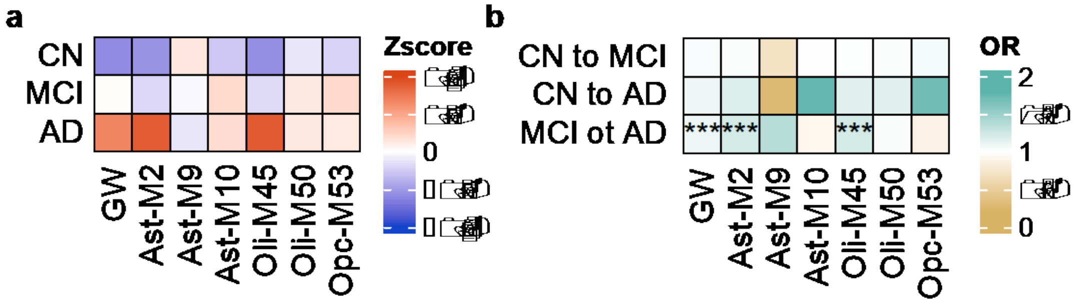

**Supplementary Figure 6.**
Association of quantitative cbPRSs and genome-wide AD PRS (gwPRS) with previously defined subgroups in ADNI 1-2/GO. PRS association with cognitively defined subgroups include age at last as a covariate. Association of PRS with MRI atrophy subtypes include age at transition as a covariate.

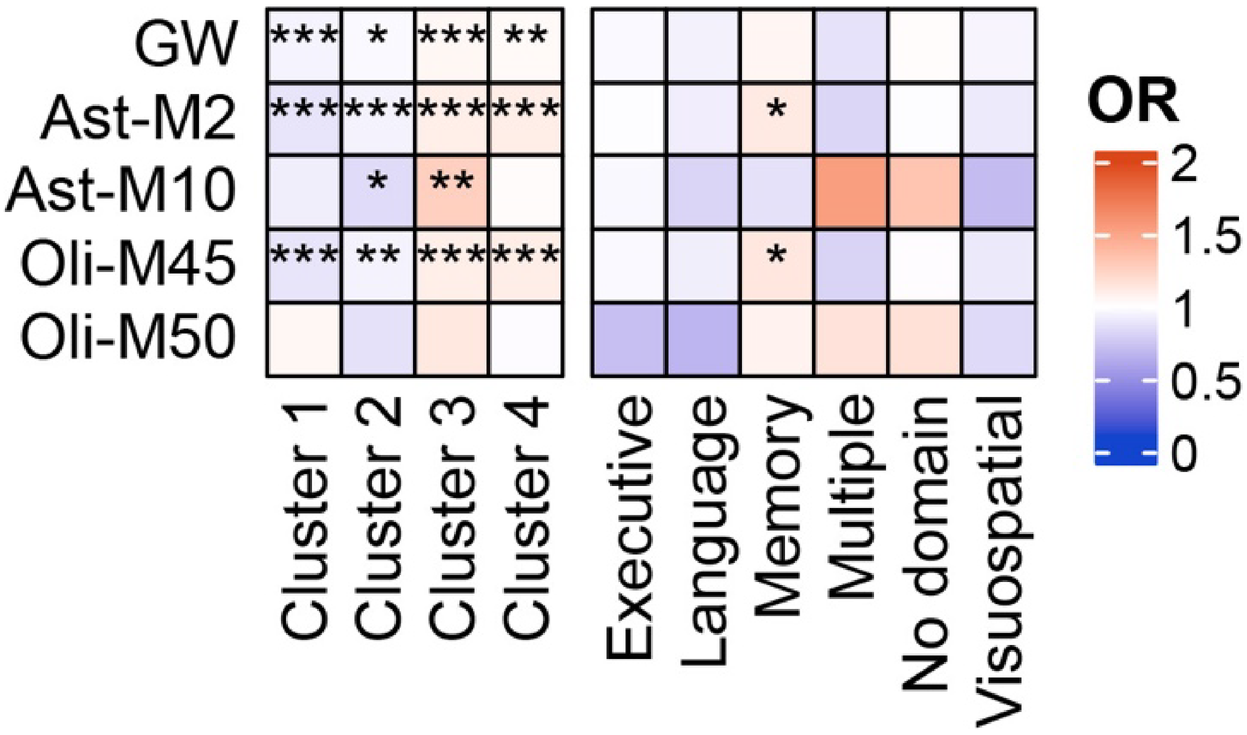

**Supplementary Figure 7.**
Association of quantitative cbPRSs with previously defined subgroups in ADNI 1-2/GO. **a.** Association of cbPRS with cognitively defined subgroups including age at last as a covariate. **b.** Association of cbPRS with MRI imaging clusters including age at transition additional covariate. Cognitive associations are not survived after multiple test correction with a Bonferroni corrected significance of 8.3E-4.

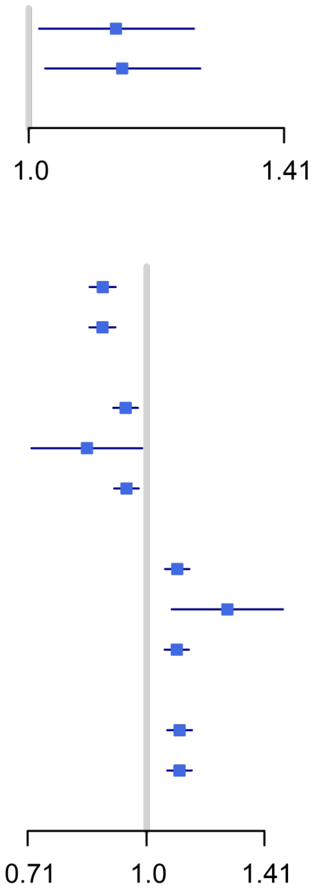

**a**

| Cognitive Domain | Module | OR[95%CI] | P |
|---|---|---|---|
| Memory | Ast-M2 | 1.12 [1.01−1.25] | 2.7e−02 |
| | Oli-M45 | 1.13 [1.02−1.26] | 1.8e−02 |

**b**

| ADNI Image | Module | OR[95%CI] | P |
|---|---|---|---|
| 1 | Ast-M2 | 0.88 [0.85−0.91] | 4.0e−11 |
| | Oli-M45 | 0.88 [0.85−0.91] | 2.5e−11 |
| 2 | Ast-M2 | 0.94 [0.91−0.98] | 8.6e−04 |
| | Ast-M10 | 0.84 [0.71−0.99] | 3.5e−02 |
| | Oli-M45 | 0.94 [0.91−0.98] | 1.3e−03 |
| 3 | Ast-M2 | 1.09 [1.06−1.13] | 7.2e−07 |
| | Ast-M10 | 1.27 [1.08−1.49] | 4.4e−03 |
| | Oli-M45 | 1.09 [1.05−1.13] | 9.7e−07 |
| 4 | Ast-M2 | 1.10 [1.06−1.14] | 1.6e−07 |
| | Oli-M45 | 1.10 [1.06−1.14] | 1.6e−07 |

**Supplementary Figure 8.**
Expression profiles of the PageRank prioritized genes in Ast-M2 perturbed by enriched drugs. Drugs shown perturb APOE expression. Genes are among the top 10 in Ast-M2. Profiles display cell-specific expression and highlight distinct biological pathways for priority genes. Genes were filtered by differential expression at the cell with nominal significance.

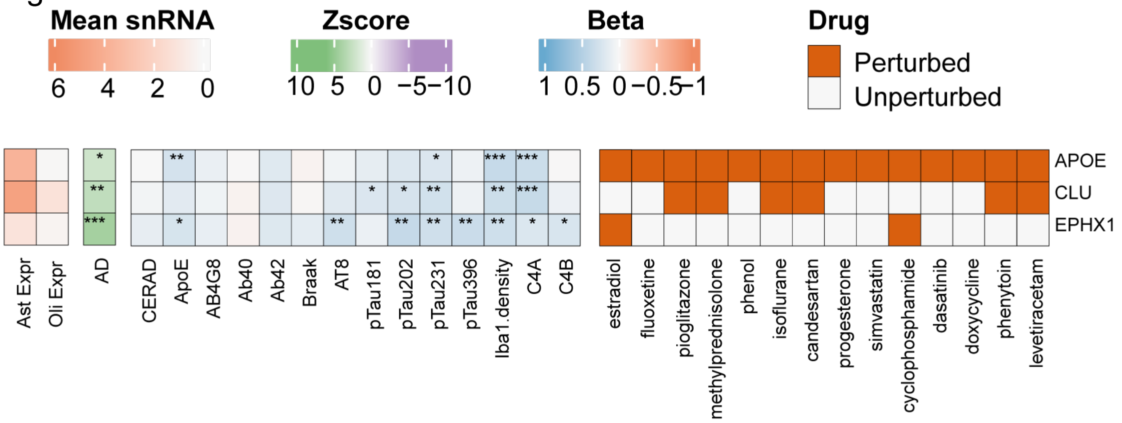

**Supplementary Figure 9.**
Summary of relative expression levels of *C4a*, *C4b* and *APOE* in heatmap. The clustering indicates similar expression patterns across samples.

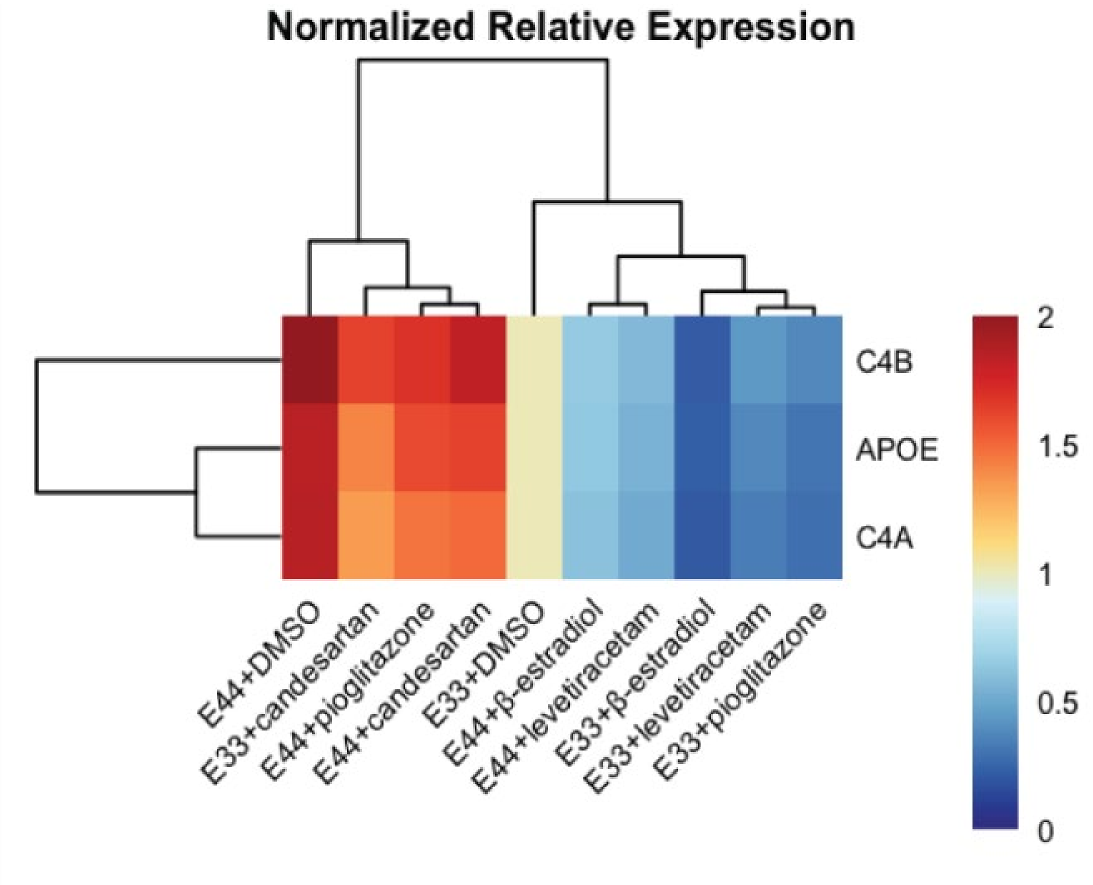

**Supplementary Figure 10.**

Measures of drug treatment efficacy in lowering *C4a* and *C4b*. expression in *APOE*33 and *APOE*44 human iPSC-derived astrocytes.

a

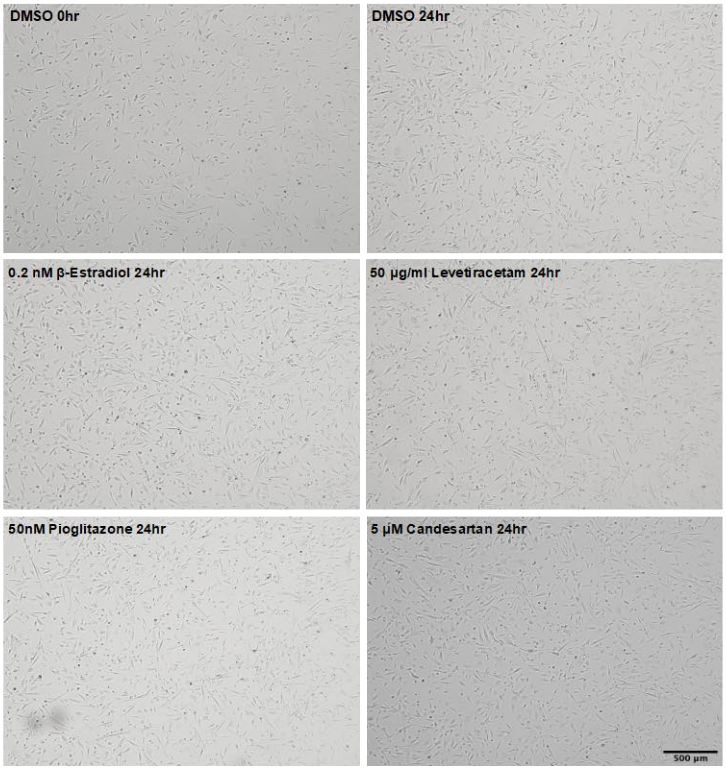

b

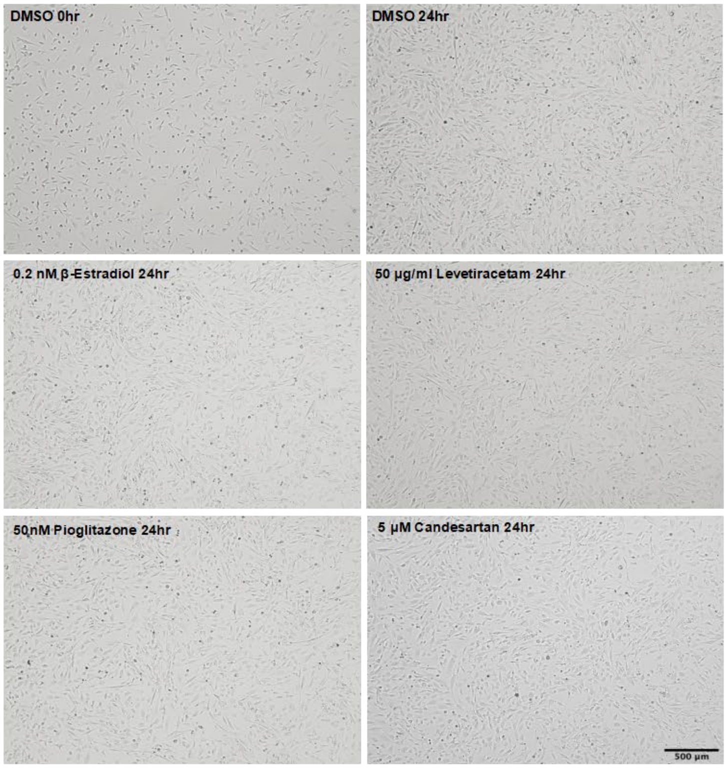

**Supplementary Figure 11.**

Measures of drug treatment efficacy in lowering C4A and C4B expression in APOE 33 and APOE 44 human iPSC-derived astrocytes. Relative expression of C4A, C4B and APOE genes in APOE 33 (E33) and APOE 44 (E44) iPSC-derived astrocytes treated with β-estradiol, **a**, pioglitazone or **b**, candesartan, respectively. Gene expression was evaluated by RT-qPCR and presented as relative fold change over the DMSO vehicle in APOE 33 astrocytes. Error bar represented the standard error of the mean. Statistics: n=3, N=3 iPSC lines per genotype; one-tailed student t-test, *p≤0.05, **p≤0.01, ***p≤0.001, ****p≤0.0001. e, Summary of relative expression levels of C4A, C4B and APOE in heatmap. The clustering indicates similar expression patterns across samples.

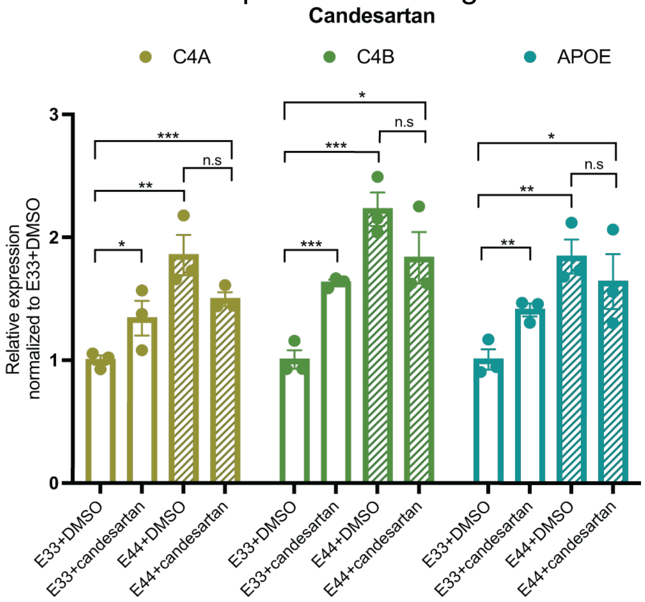

### References

1. Kunkle, B.W., *et al.* Genetic meta-analysis of diagnosed Alzheimer's disease identifies new risk loci and implicates Aβ, tau, immunity and lipid processing. *Nature Genetics* **51**, 414-430 (2019).
